# Heart rate rises most during REM sleep after semaglutide initiation: a matched longitudinal cohort study

**DOI:** 10.64898/2026.10.18.26365823

**Authors:** David Krongauz, Sarah Kohn, Arad Zulti, Yishay Schlesinger, Anastasia Godneva, Yeela Talmor-Barkan, Eran Segal

**Author notes:** Corresponding author: Eran Segal, Department of Computer Science and Applied Mathematics, Weizmann Institute of Science, 234 Herzl Street, Rehovot 7610001, Israel.

## Abstract

**Study Objectives.:** Glucagon-like peptide-1 (GLP-1) receptor agonists raise resting heart rate. We asked whether, after semaglutide initiation, the increase is larger during rapid eye movement (REM) sleep than during non-rapid eye movement (NREM) sleep.

**Methods.:** In the Human Phenotype Project, an Israeli cohort of generally healthy adults aged 40 to 70 years, 96 of 494 self-reported semaglutide initiators had a home sleep study (WatchPAT; sleep stages estimated from peripheral arterial tone and actigraphy) before and at least three months after starting. Each was propensity-matched 1:1 to a never-user of GLP-1 drugs measured at the same visits. We estimated the initiators’ change minus their controls’ change (difference-in-differences). The REM minus NREM contrast was designated primary after an exploratory analysis.

**Results.:** Relative to controls (90 to 94 pairs), heart rate rose during nocturnal wake (+3.89 beats per minute [bpm], 95% confidence interval 2.35 to 5.44), NREM sleep (+4.82 bpm, 3.12 to 6.52) and REM sleep (+6.25 bpm, 4.42 to 7.91); the REM increase exceeded the NREM increase by 1.42 bpm (0.98 to 1.89). The excess persisted across matching designs and on a relative scale; the proportion of REM sleep did not change. Never-users with similar weight loss showed no REM sleep heart-rate rise (+0.40 bpm, −0.63 to 1.50). Heart-rate variability decreases disappeared after adjustment for heart rate.

**Conclusions.:** Semaglutide initiation was associated with higher heart rate throughout sleep, most during REM sleep, unexplained by weight loss. Clinic measurements may understate this effect. Confirmation with polysomnographic staging is needed.

**Graphical Abstract:** 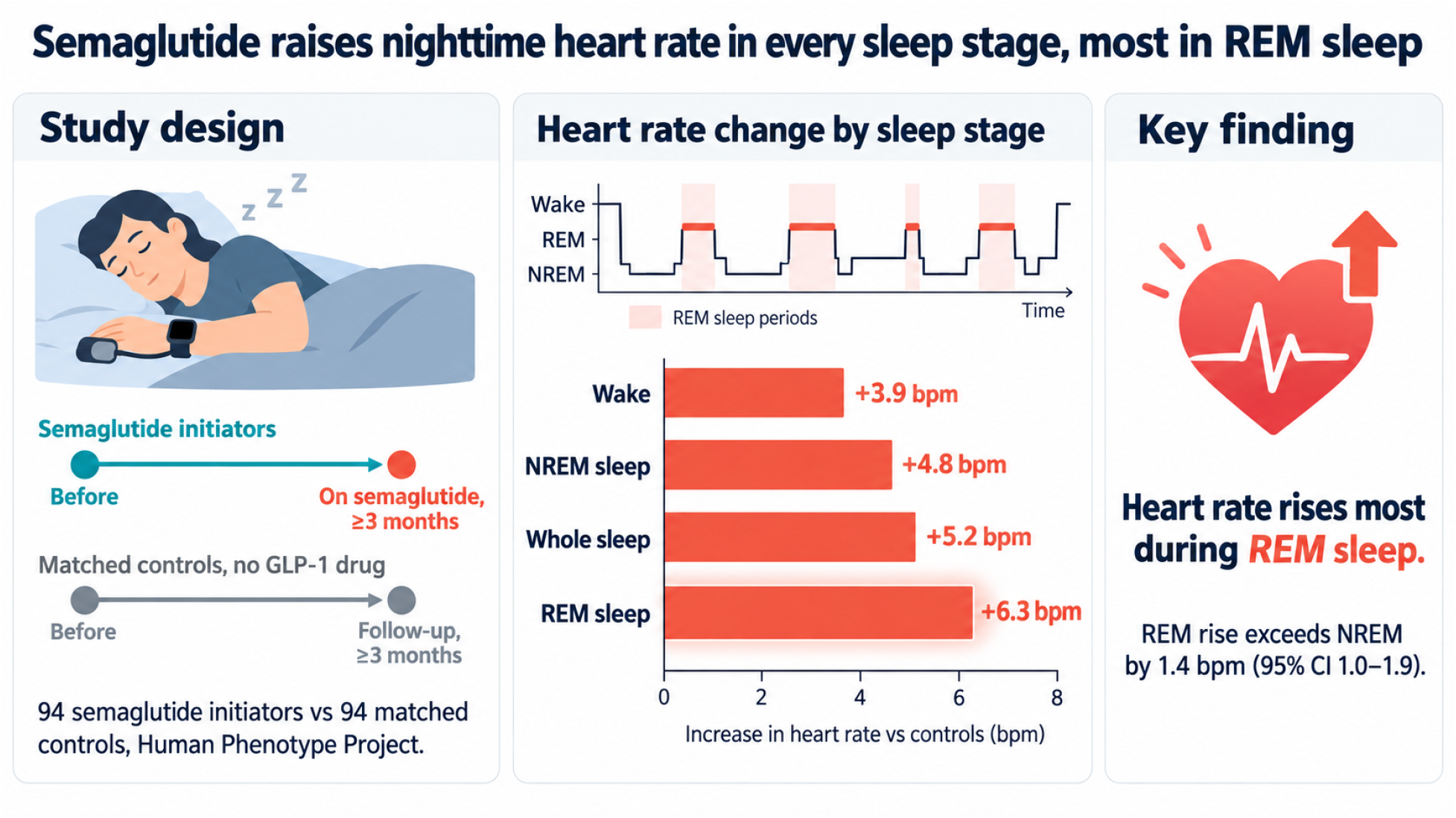

**Statement of Significance:** Glucagon-like peptide-1 receptor agonists lower body weight and cardiovascular risk but raise heart rate, an effect usually measured while patients sit awake in a clinic. Whether it differs between sleep stages has not been formally tested. After people started semaglutide, heart rate rose in every sleep stage estimated by a home sleep apnea test, most during rapid eye movement sleep; weight loss did not explain it. A higher sleeping heart rate on a wearable after starting the drug may therefore be an expected effect, not illness. Whether the rise matters for patients with heart disease, in whom rapid eye movement sleep can provoke ischemia and arrhythmia, requires prospective studies with dispensed exposure and electroencephalographic sleep staging.

## Introduction

Glucagon-like peptide-1 (GLP-1) receptor agonists have become central to the treatment of obesity and cardiometabolic disease. Randomized trials of semaglutide show substantial weight loss and cardiovascular benefit [1–3]. Alongside these benefits, GLP-1 receptor agonists consistently increase resting heart rate [4,5]. The increase is usually modest and has not offset the cardiovascular benefit observed in outcome trials [2], but it runs against the direction in which resting heart rate is otherwise associated with cardiovascular and all-cause mortality [6,7], and its physiological origin and timing across the day remain incompletely resolved. Candidate mechanisms include a direct action at the sinoatrial node and central autonomic modulation, both supported by receptor distribution but not resolved in humans [8].

Continuous and ambulatory studies have extended the trial observation. Liraglutide increased mean and nighttime heart rate in a randomized crossover study, with accompanying changes in some heart-rate-variability (HRV) measurements [9]. A recent wearable study similarly reported increased resting heart rate after GLP-1 receptor agonist initiation [10]. These studies establish the heart-rate increase, but they do not formally compare it between rapid eye movement (REM) sleep, non-rapid eye movement (NREM) sleep and wake.

Sleep offers a useful physiological comparison because autonomic and cardiovascular regulation differ substantially between sleep stages. Muscle sympathetic nerve activity falls progressively through NREM sleep and rises during REM sleep to levels exceeding quiet wakefulness, with surges in heart rate and blood pressure [11]; cardiac autonomic balance follows the same stage sequence across the night [12]. REM sleep is therefore the stage in which heart rate is most variable and least tightly regulated, so a drug that shifts heart rate may express that shift differently there than during NREM sleep. Resolving a drug-associated heart-rate effect by sleep stage may help locate when the effect is most expressed, even when an observational study cannot identify its cause.

The Human Phenotype Project (HPP) combines repeated medication reconciliation with anthropometrics, blood pressure, body composition, multi-night home sleep monitoring, diet, blood tests and gut metagenomics [13,14]. We used this longitudinal depth to emulate a trial of semaglutide initiation as closely as observational data allow, with a matched difference-in-differences design, in which each measurement domain had to pass prespecified balance checks before its results were interpreted. The central question was whether the accompanying heart-rate increase was uniform across sleep stages or larger during REM sleep. Secondary analyses tested alternative matching designs, redrew the match within a bootstrap, compared exposure definitions, examined whether the larger REM increase could reflect proportional scaling, baseline heart rate, weight change or the device’s stage estimation, and asked whether apparent HRV changes persisted after adjustment for heart rate. A phenome-wide analysis of the other measurement domains tests whether the design recovers the effects already established in randomized trials, which supports its validity, rather than adding headline claims.

## Methods

### Study population and exposure

The HPP is a longitudinal cohort of adults with repeated clinical, behavioral, physiological and molecular measurements at study visits scheduled every two years [13]. Recruitment began in Israel in 2018 and enrolls generally healthy adults aged 40 to 70 years; exclusion criteria at enrollment include cardiovascular, chronic respiratory, kidney, gastrointestinal, neurological or metabolic disease (including diabetes), malignancy and pregnancy [13]. The visits analyzed here took place between 2019 and 2026, and the sleep studies between 2020 and 2025. At each visit participants report their current medications with start dates. Initiators were participants with a semaglutide record, under its generic or any brand name, carrying a plausible start date (2018 or later) and no record of any other GLP-1 receptor agonist, including tirzepatide, anywhere in their medication history (n=494). Controls were participants with no GLP-1 receptor agonist record of any kind; the 788 participants with any such record were excluded from the control pool.

The HPP protocol was approved by the Institutional Review Board of the Weizmann Institute of Science (approval no. 2392-4), and all participants provided written informed consent. The study was conducted in accordance with the Declaration of Helsinki. Methods and results are reported following the Strengthening the Reporting of Observational Studies in Epidemiology (STROBE) statement.

### Home sleep monitoring

Sleep was recorded at home with the WatchPAT-300 (Itamar Medical), a home sleep apnea test that combines a wrist actigraph, a finger probe measuring peripheral arterial tone (PAT) and pulse oximetry, and a chest sensor for snoring and body position. At each visit participants recorded up to three nights within two weeks. The device algorithm estimates wake, light sleep (N1 and N2), deep sleep (N3) and REM sleep from PAT, pulse rate and actigraphy; it does not record electroencephalography. Its respiratory indices agree closely with polysomnography [15], and its sleep-stage estimates have been validated against polysomnography [16,17]. The device algorithm detects respiratory events from PAT amplitude, pulse rate and oxygen saturation: the AHI counts hypopneas with at least 3% oxygen desaturation, the 4%-desaturation AHI those with at least 4%, and the ODI counts desaturations of at least 4% per hour of sleep. For each night we used the mean heart rate in each sleep stage, and during wake periods within the recording (nocturnal wake), when at least five minutes were spent in it and the heart-rate signal-quality score was at least 50, and we averaged the nights of each visit. Sleep outcomes were heart rate during sleep overall (the mean across all sleep stages, excluding wake), nocturnal wake, NREM sleep and REM sleep; the apnea-hypopnea index (AHI), 4%-desaturation AHI and oxygen desaturation index (ODI), and the AHI, ODI and respiratory disturbance index (RDI) during REM sleep; sleep efficiency; and time-domain HRV (root mean square of successive differences [RMSSD], standard deviation of inter-beat intervals [SDNN] and proportion of successive intervals differing by more than 20 ms [pNN20]) during REM and NREM sleep.

### Modalities and outcomes

We call each measurement domain a modality: sleep, anthropometrics, blood pressure, dual-energy x-ray absorptiometry (DXA) body composition, blood tests, gut taxonomy, gut microbiome function and diet. Weight, height, waist and hip circumferences, sitting blood pressure and pulse, and whole-body DXA (Lunar Prodigy Advance with CoreScan, GE Healthcare, which also estimates visceral adipose tissue area) were measured at study visits; blood tests were taken from participants’ health maintenance organization records; stool was self-collected with OMNIgene·GUT kits (DNA Genotek) and shotgun-sequenced; and diet was logged in a smartphone app for up to two weeks [13]. Protocols are detailed in the Supplementary Methods. Each modality contains several outcomes, the individual measured variables, such as REM sleep heart rate within the sleep modality. Microbial pathways present in at least 10% of samples were centered log-ratio transformed, leaving 272 functional pathways; 321 outcomes were tested in total.

### Visit selection and matching

For each initiator and modality, the baseline visit was the last visit with a usable measurement before the recorded start date, and the follow-up visit the first usable visit at least three months after it. A three-month minimum places the follow-up visit after the standard semaglutide dose escalation for diabetes (0.25 to 0.5 to 1.0 mg over about eight weeks), although escalation for obesity (to 2.4 mg over 16 weeks) may still be under way at three months.

Matching was done once for each modality, because the participants measured at both visits differ between modalities, and it used only baseline covariates, never outcomes; within a modality, the same matched pairs serve all its outcomes. The propensity model included age, body-mass index (BMI), waist and hip circumference, visceral adipose tissue area (with an indicator for a missing measurement), sex, diabetes, current smoking, antihypertensive therapy and lipid-lowering therapy at the baseline visit; diabetes, smoking and medication use were taken from participant self-report. Each initiator was matched to one control without replacement, among controls with the same baseline and follow-up visits, the same sex and the same lipid-lowering status whose propensity logit lay within 0.2 standard deviations; among these, the assignment minimized the total Mahalanobis distance. A modality was interpreted only if matching retained at least 85% of eligible initiators and every covariate had a post-match absolute standardized mean difference (SMD) below 0.10. These matching criteria were checked before any outcome was examined. Outcomes, including pre-treatment heart rate, were not matched on: matching on a pre-treatment outcome selects controls from the tail of their distribution, whose regression toward the mean then biases the difference-in-differences in the direction of the treated group [18].

### Estimation and multiplicity

For each pair, the difference-in-differences (DiD) was the initiator’s change from the baseline to the follow-up visit minus the matched control’s change. The estimate is the mean DiD, with a 95% confidence interval (CI) from 2,000 bootstrap resamples of pairs. Each outcome was analyzed in the pairs in which both members had a measurement at both visits (complete-case analysis), so the number of pairs differs between outcomes and is reported with each estimate (Supplementary Table S1). Participants missing age, sex, BMI, or waist or hip circumference could not be matched; visceral adipose tissue area, the only other covariate with missing values, was imputed and flagged with a missing-measurement indicator (Supplementary Methods). No sample size was calculated in advance: every eligible initiator was included, and the precision of each estimate is given by its CI. Heart-rate contrasts between sleep stages and states (REM minus NREM sleep, REM sleep minus nocturnal wake, NREM sleep minus nocturnal wake) were estimated within pairs on the pairs with both values. The contrasts with nocturnal wake, the step in heart rate at REM onset and the change in the proportion of sleep estimated as REM sleep have paired t intervals.

False discoveries were controlled hierarchically [19,20]: Benjamini-Hochberg correction within each modality, and a Simes test for each modality with Benjamini-Hochberg correction across the modalities that passed their matching criteria. An outcome was a discovery if its modality was selected and its within-modality q was below 0.05. Modalities that failed a matching criterion are reported but could neither contribute discoveries nor enter the correction. Under the stricter selective-inference procedure of Benjamini and Bogomolov [21], which tests within the three selected of five eligible modalities at 0.05×3/5, 19 of the 23 discoveries remain; diastolic blood pressure (q=0.043), the 4%-desaturation AHI (q=0.031), ODI (q=0.039) and REM sleep AHI (q=0.048) do not. REM sleep heart rate was designated the primary sleep outcome, and the REM minus NREM heart-rate difference the primary contrast, after an earlier exploratory analysis of these data; they are therefore internally validated rather than prospectively confirmed.

### Sensitivity and interpretation analyses

Two alternative designs were estimated: variable 1:1 to 1:3 matching in which no control was used more than twice, with heteroskedasticity-consistent (HC3) standard errors; and overlap weighting within each exact-matching stratum [22], which balances every covariate in the propensity model by construction, with participant-clustered standard errors. To carry the uncertainty of the matching step into the interval, 500 bootstrap replicates resampled initiators, refit the propensity model and redrew the match. Two alternative exposure definitions (starters of any GLP-1 receptor agonist, and starters of liraglutide, dulaglutide or tirzepatide) were analyzed with the same procedure.

Three analyses addressed alternative explanations of a larger heart-rate increase during REM sleep. The relative-scale contrast compared the log-scale heart-rate change in REM and NREM sleep within pairs, so that a single proportional rise gives a ratio of exactly 1.0. Dependence on baseline heart rate was estimated by regressing each DiD on the initiator’s mean of baseline and follow-up heart rate (Oldham’s method), which avoids the artifact that regression to the mean creates when a change is regressed on its own starting value [23]. The REM minus NREM heart-rate difference was regressed on the nocturnal wake DiD, whose intercept estimates the REM minus NREM difference at zero heart-rate change during nocturnal wake. HRV outcomes were adjusted for the simultaneous heart-rate DiD in the same sleep stage [24], and REM sleep heart rate for the pair’s weight change, by regression with HC3 standard errors; the adjusted intercept is descriptive, not a mediation estimate. On the same pairs, the REM minus NREM heart-rate difference was regressed on each pair’s change in the proportion of sleep estimated as REM sleep, and each DiD on the initiator’s months from the recorded start to the follow-up visit, with HC3 standard errors. Sleep-stage proportions across all HPP visits with more than three hours of sleep were compared with polysomnographic norms. The change in matched controls was compared with the change in all eligible participants who never took a GLP-1 drug over the same visit intervals. Post hoc subgroup analyses repeated the REM sleep heart-rate and REM minus NREM estimates by the initiator’s sex, age, BMI and antihypertensive therapy at baseline; the difference between two subgroups was estimated by regressing the pair DiD on the subgroup indicator, with HC3 standard errors. To ask whether weight loss itself raises heart rate during sleep, we related each participant’s change in sleep heart rate to their change in weight and in DXA fat mass among participants who never took a GLP-1 drug (each person’s earliest pair of consecutive visits with a sleep study and weight at both), and among initiators, by linear regression adjusted for age, sex, baseline BMI and the interval between visits, with HC3 standard errors. Each initiator was then matched to one such participant, without reuse, by the primary matching procedure with the size of the weight or DXA fat loss and the interval between visits added to the propensity model; the difference in heart-rate change has a 95% CI from 2,000 bootstrap resamples of pairs. Finally, the device’s per-second recordings of heart rate and sleep stage, which resolve NREM sleep into light and deep sleep, were matched by the same procedure within the primary sleep cohort. Analyses were performed in Python 3.11; the package versions are listed in the code repository (Code availability).

## Results

### Study cohort

The HPP measures the same participants repeatedly over years, including home sleep monitoring, so a change in heart rate after starting a drug can be compared with the change in similar people who did not start it. Of the 494 semaglutide initiators, 96 had a sleep study before the recorded start and another at least three months after it, and 94 were matched to 94 controls who never took a GLP-1 drug (Figure 1a; Supplementary Figure S1). The sleep match retained 98.9% of eligible initiators with a maximum post-match SMD of 0.089 (Table 1; Figure 2c). The matched groups were 50% female, with a mean age of 53 years and a mean BMI of 32 kg/m². The follow-up visit came a median of 13.1 months after the recorded start (range 3.1 to 49.9), and the median interval between visits was 24.8 months in initiators and 25.3 months in controls. Although heart rate was not a matching variable, pre-treatment sleep heart rate differed between groups by at most 0.96 beats per minute (bpm) (nocturnal wake; SMD at most 0.12).

**Figure 1.**
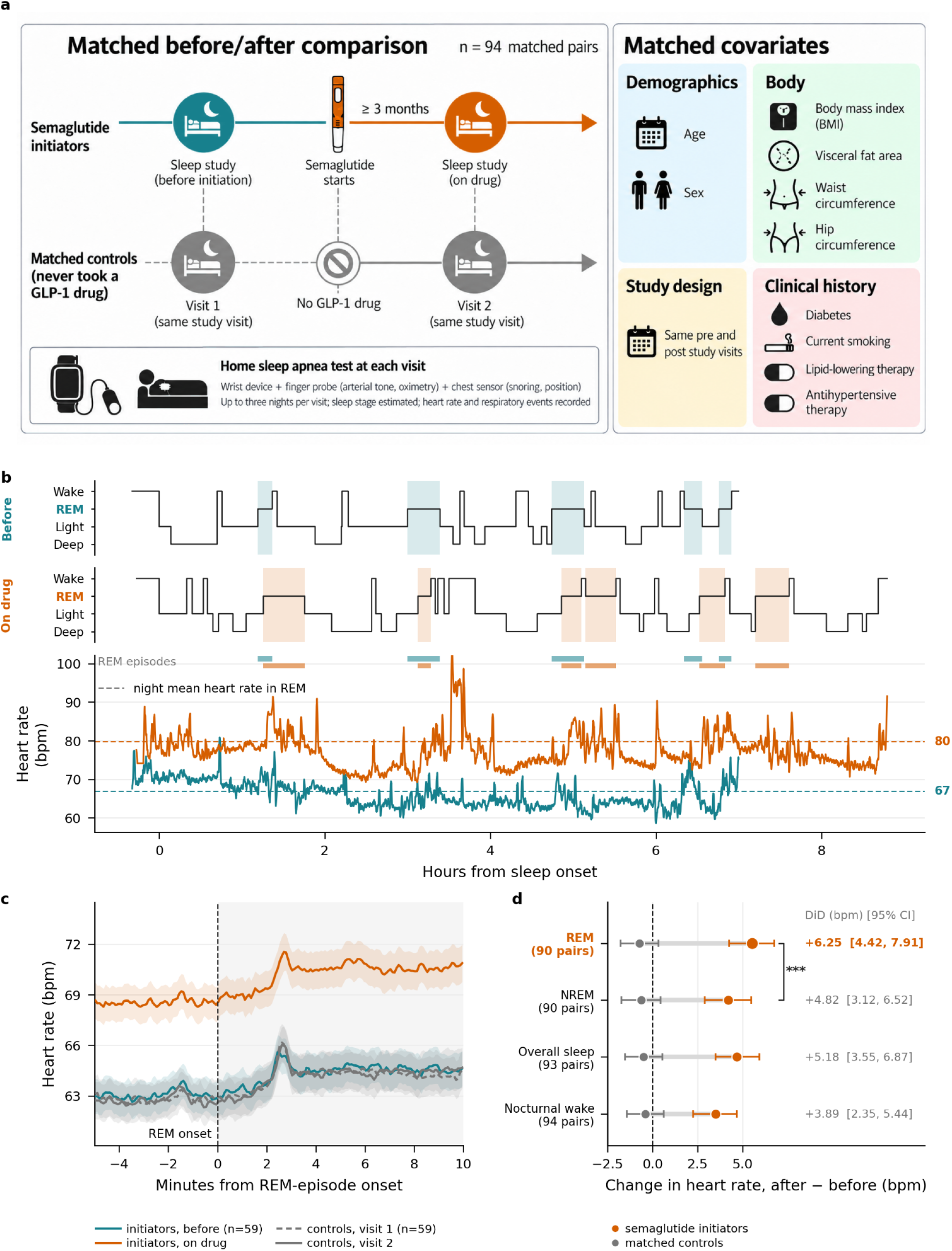
Home sleep monitoring shows the semaglutide-associated heart-rate increase is largest during REM sleep. (a) Study design and matched covariates; teal, before initiation; vermilion, on drug; gray, matched controls. (b) One illustrative initiator’s hypnograms before initiation (teal) and on drug (vermilion), with REM episodes shaded in the same colors, above the heart-rate traces; dashed lines mark each night’s mean heart rate during REM sleep. (c) Mean heart rate around the onset of REM episodes of at least 3 minutes in 59 matched pairs from the per-second recordings; initiators before initiation (teal) and on drug (vermilion), controls at visit 1 (gray dashed) and visit 2 (gray solid); bands, ±1 standard error of the mean (SEM). (d) Change in heart rate from baseline to follow-up in initiators (vermilion) and controls (gray) by sleep stage and state, with 95% CIs; numbers at right are the DiD (95% CI). ***q<0.001 for the REM minus NREM heart-rate contrast. *Alt text: Four-panel figure. Panel a: a schematic of the matched design, with a teal before-initiation sleep study and a vermilion on-drug sleep study for initiators, gray studies for matched controls, and icons for the matched covariates. Panel b: one participant’s two hypnograms, with REM episodes shaded teal before initiation and vermilion on drug, above heart-rate traces in the same colors, with each night’s mean REM heart rate as a dashed line, 67 bpm before and 80 bpm on drug. Panel c: cohort mean heart rate around REM onset; the initiators’ on-drug curve in vermilion lies about 6 bpm above their before curve in teal, which overlaps the two gray control curves, and every curve peaks about 3 bpm above its pre-onset level between 2 and 3 minutes after onset. Panel d: a dumbbell plot of four sleep stages and states, with the DiD at right: REM sleep +6.25 bpm, NREM sleep +4.82, sleep overall +5.18 and nocturnal wake +3.89, every DiD interval excluding zero, and a bracket marking the REM minus NREM contrast*.

**Figure 2.**
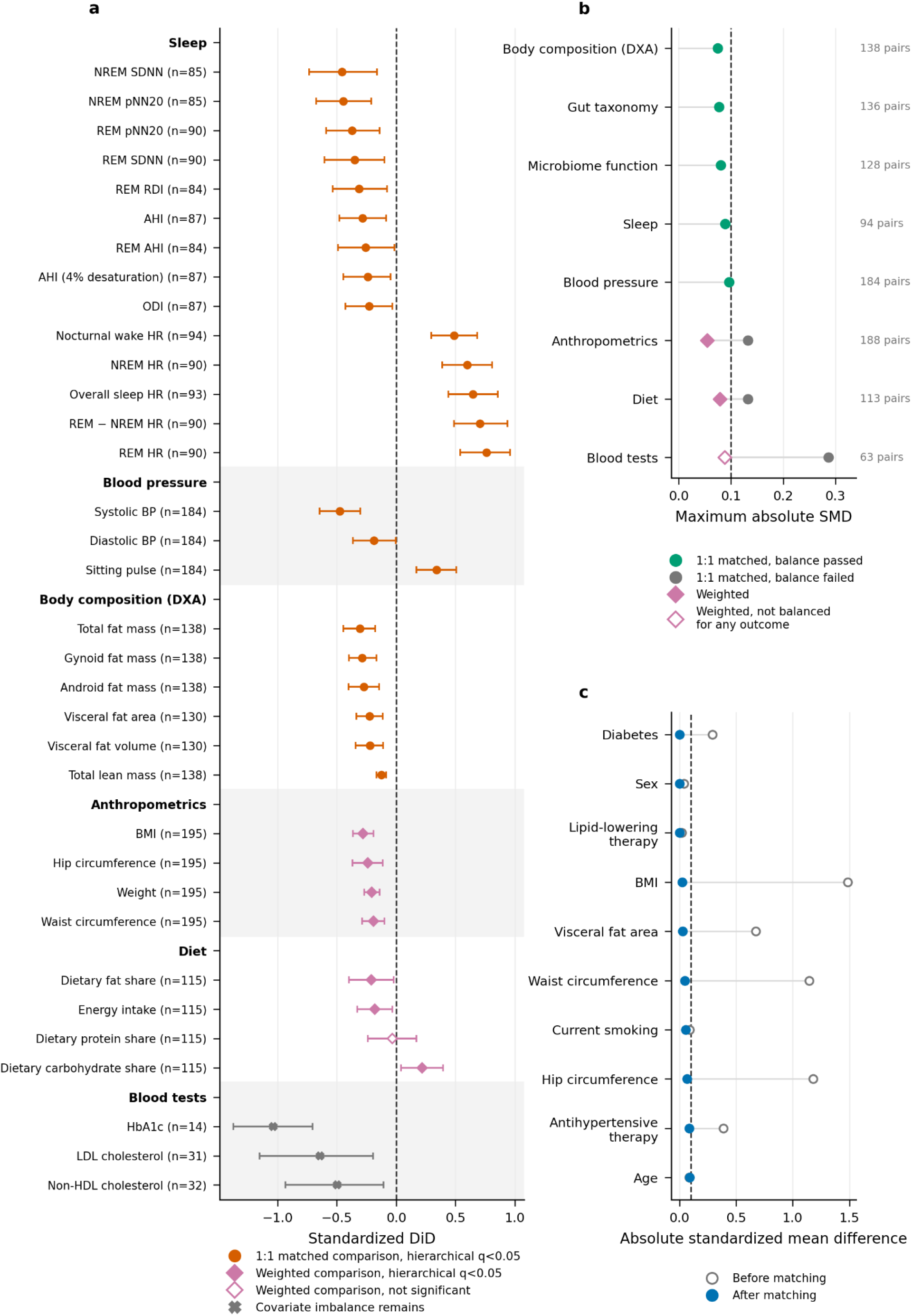
Clinical and multi-omic findings. (a) Standardized DiD (effect divided by the baseline SD) with 95% CIs. Vermilion circles, discoveries from the 1:1 match; pink diamonds, overlap-weighted estimates for modalities whose 1:1 match failed balance (filled, passed the false-discovery procedure; open, did not); gray crosses, blood-test estimates that remained imbalanced. Row labels give pairs (1:1) or initiators (weighted). (b) Maximum absolute SMD by modality under 1:1 matching (green, passed; gray, failed) and overlap weighting (pink; open, not balanced for any outcome); dashed line, 0.10; labels, matched pairs. (c) Sleep-match covariate balance before (open circles) and after (blue) matching. HR, heart rate; BP, blood pressure. *Alt text: Three-panel figure. Panel a is a forest plot of 34 outcomes in baseline standard deviations, grouped as sleep, blood pressure, DXA body composition, anthropometrics, diet and blood tests. Vermilion circles mark the 23 discoveries from balanced 1:1 matches: sleep heart rate rises, HRV and respiratory indices fall, blood pressure and DXA fat and lean mass fall, and sitting pulse rises. Pink diamonds mark weighted estimates; the filled ones are all four anthropometric outcomes, energy intake and the dietary fat and carbohydrate shares. Gray crosses mark three blood-test estimates. Panel b shows maximum absolute SMD per modality against a dashed line at 0.10, green for the five modalities that passed and gray for the three that failed, each joined to a pink diamond for its weighted analysis; the blood-test diamond is open because the weighted analysis was not balanced for any outcome. Panel c shows covariate balance before matching as open circles and after matching as blue circles; every post-match value is below 0.10*.

**Table 1.**
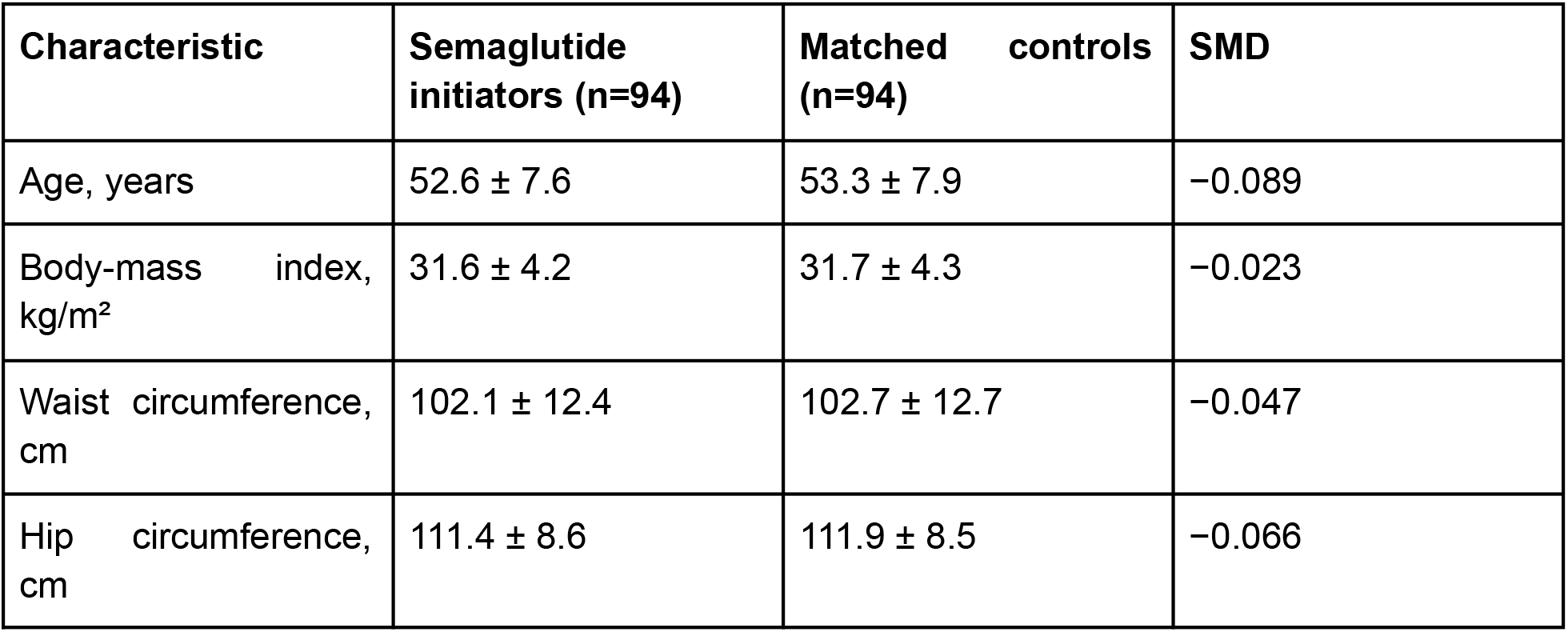
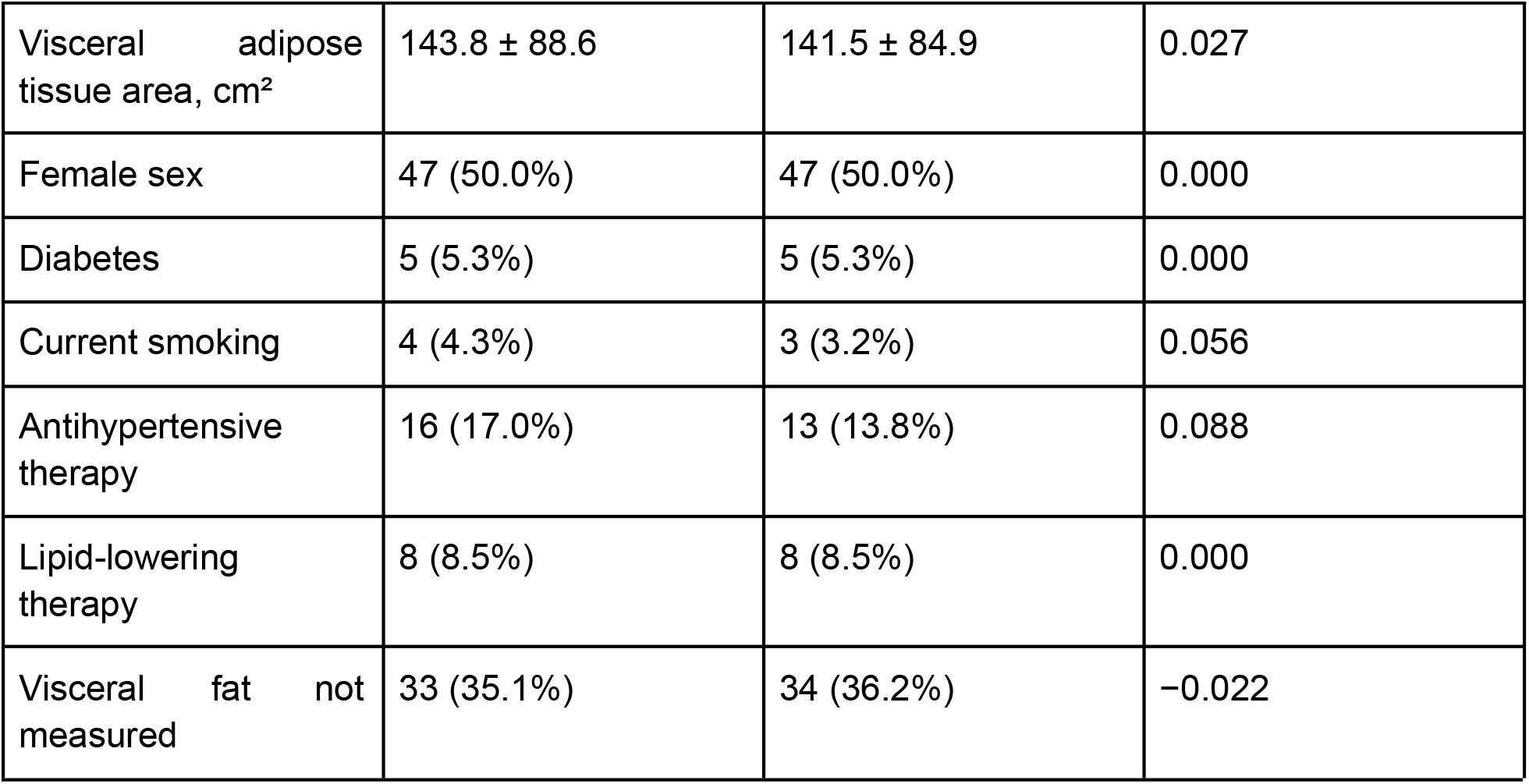
Baseline characteristics of the matched sleep cohort. Values at the baseline visit. Continuous variables are mean ± standard deviation (SD) and binary variables n (%). SMD is the post-match standardized mean difference. Sex and lipid-lowering therapy were matched exactly.

| Characteristic | Semaglutide initiators (n=94) | Matched controls (n=94) | SMD |
| --- | --- | --- | --- |
| Age, years | 52.6 ± 7.6 | 53.3 ± 7.9 | -0.089 |
| Body-mass index, kg/m<sup>2</sup> | 31.6 ± 4.2 | 31.7 ± 4.3 | -0.023 |
| Waist circumference, cm | 102.1 ± 12.4 | 102.7 ± 12.7 | -0.047 |
| Hip circumference, cm | 111.4 ± 8.6 | 111.9 ± 8.5 | -0.066 |
| Visceral adipose tissue area, cm<sup>2</sup> | 143.8 ± 88.6 | 141.5 ± 84.9 | 0.027 |
| Female sex | 47 (50.0%) | 47 (50.0%) | 0.000 |
| Diabetes | 5 (5.3%) | 5 (5.3%) | 0.000 |
| Current smoking | 4 (4.3%) | 3 (3.2%) | 0.056 |
| Antihypertensive therapy | 16 (17.0%) | 13 (13.8%) | 0.088 |
| Lipid-lowering therapy | 8 (8.5%) | 8 (8.5%) | 0.000 |
| Visceral fat not measured | 33 (35.1%) | 34 (36.2%) | -0.022 |

Of the eight modalities, sleep, blood pressure, DXA, gut taxonomy and microbiome function passed both matching criteria, and anthropometrics, blood tests and diet failed the balance check (Figure 2b; Supplementary Table S2).

### Heart rate rises in every sleep stage and most during REM sleep

Among the matched pairs (90 to 94 per outcome, depending on complete data), heart rate rose during sleep overall (+5.18 bpm, 95% CI 3.55 to 6.87), nocturnal wake (+3.89 bpm, 2.35 to 5.44), NREM sleep (+4.82 bpm, 3.12 to 6.52) and REM sleep (+6.25 bpm, 4.42 to 7.91) (Figure 1d; Table 2). The increase came from the initiators: REM sleep heart rate rose by 5.52 bpm in initiators and fell by 0.72 bpm in their controls. In the 90 pairs with both sleep stages, the heart-rate increase during REM sleep exceeded that of NREM sleep by 1.42 bpm (0.98 to 1.89; q=4.1×10⁻⁷) and that of nocturnal wake by 2.30 bpm (1.42 to 3.18); the increase during NREM sleep also exceeded that of nocturnal wake, by 0.87 bpm (0.11 to 1.64). Heart rate therefore rose most during REM sleep, less during NREM sleep and least during nocturnal wake.

**Table 2.** Primary estimates in the modalities that passed their matching criteria. DiD with 95% bootstrap CIs; q is the Benjamini-Hochberg value within the modality. AHI, apnea-hypopnea index; BP, blood pressure; bpm, beats per minute; CI, confidence interval; DiD, difference-in-differences; ODI, oxygen desaturation index.

| Outcome | Unit | Pairs | DiD (95% CI) | $q$ (within modality) | Discovery |
| --- | --- | --- | --- | --- | --- |
| Heart rate, sleep overall | bpm | 93 | +5.18 (3.55 to 6.87) | $2.5 \times 10^{-7}$ | yes |
| Heart rate, nocturnal wake | bpm | 94 | +3.89 (2.35 to 5.44) | $1.9 \times 10^{-5}$ | yes |
| Heart rate, NREM sleep | bpm | 90 | +4.82 (3.12 to 6.52) | $1.2 \times 10^{-6}$ | yes |

| Outcome | Unit | Pairs | DiD (95% CI) | q (within modality) | Discovery |
| --- | --- | --- | --- | --- | --- |
| Heart rate, REM sleep | bpm | 90 | +6.25 (4.42 to 7.91) | $2.9 \times 10^{-9}$ | yes |
| Heart rate, REM minus NREM sleep | bpm | 90 | +1.42 (0.98 to 1.89) | $4.1 \times 10^{-7}$ | yes |
| Apnea-hypopnea index (AHI) | events/h | 87 | -3.93 (-6.64 to -1.15) | 0.014 | yes |
| Oxygen desaturation index (ODI) | events/h | 87 | -2.57 (-4.84 to -0.36) | 0.039 | yes |
| Systolic BP | mmHg | 184 | -7.51 (-10.21 to -4.77) | $5.9 \times 10^{-7}$ | yes |
| Diastolic BP | mmHg | 184 | -2.05 (-4.00 to -0.02) | 0.043 | yes |
| Sitting pulse | bpm | 184 | +3.83 (1.89 to 5.65) | $8.9 \times 10^{-5}$ | yes |
| Total fat mass | kg | 138 | -2.35 (-3.42 to -1.35) | $2.7 \times 10^{-5}$ | yes |
| Total lean mass | kg | 138 | -1.33 (-1.74 to -0.89) | $5.9 \times 10^{-8}$ | yes |
| Visceral fat area | cm<sup>2</sup> | 130 | -18.51 (-27.59 to -9.23) | $1.8 \times 10^{-4}$ | yes |

Figure 1b shows the two nights of one initiator, chosen for illustration among those measured at least six months after the recorded start whose heart rate rose more during REM than NREM sleep, and Figure 1c the cohort’s heart rate around the onset of REM episodes in 59 matched pairs from the per-second recordings, a separate match that did not pass the balance check and is therefore descriptive. Heart rate rose at REM onset at every visit in both groups; after semaglutide the initiators’ whole curve shifted upward, while the step at REM onset changed little (+0.32 bpm, −0.17 to 0.81). This view samples only REM episodes of at least three minutes and the minutes around their onset, and it shows a smaller REM minus NREM difference than the whole-night estimate in the same pairs (+0.80 bpm, 0.19 to 1.39).

### A phenome-wide analysis recovers the expected clinical responses

Across the other modalities, the matched design recovered the expected clinical responses (Figure 2a; Table 2). In 184 pairs, systolic blood pressure fell by 7.51 mmHg (4.77 to 10.21) and diastolic blood pressure by 2.05 mmHg (0.02 to 4.00). Sitting pulse rose by 3.83 bpm (1.89 to 5.65), but mostly because the controls’ pulse fell (by 2.24 bpm) while the initiators’ rose by 1.58 bpm. On DXA (138 pairs), total fat mass fell by 2.35 kg (1.35 to 3.42) and lean mass by 1.33 kg (0.89 to 1.74), and in 130 pairs visceral fat area fell by 18.51 cm² (9.23 to 27.59). Respiratory indices improved: the AHI fell by 3.93 events/h (1.15 to 6.64), with consistent decreases in the 4%-desaturation AHI, REM sleep AHI, ODI and REM sleep respiratory disturbance index (Supplementary Table S1). The matched controls changed like the whole untreated cohort over the same visit intervals (5,401 to 7,782 participants who never took a GLP-1 drug, depending on the outcome) for nine of ten outcomes, including the declines in diastolic pressure and sitting pulse; the controls lost more lean mass than the untreated cohort, which if anything makes the lean-mass estimate conservative (Supplementary Figure S2).

Weight fell by 3.53 kg in the 1:1 match, but the anthropometric match failed the balance check (maximum SMD 0.132); under overlap weighting, which passed it, weight fell by 3.11 kg (2.14 to 4.08) in 195 initiators. Diet also failed balance under 1:1 matching; under overlap weighting in 115 initiators, energy intake fell by 91 kcal/day (17 to 166) and the fat share of calories by 1.34 percentage points (0.14 to 2.54), while the carbohydrate share rose by 1.80 points (0.35 to 3.25). These are post hoc estimates and are reported as hypothesis-generating. Blood tests remained imbalanced under both designs, so no blood-test result is interpreted (Figure 2b; Supplementary Figure S3). The five modalities that passed their matching criteria produced 23 discoveries: 14 in sleep, three in blood pressure and six in DXA, while gut taxonomy and microbiome function produced none (Supplementary Table S2).

### The larger REM increase is robust to design, exposure definition and scale

The larger heart-rate increase during REM sleep did not depend on the matching design. Under 1:1 to 1:3 matching with limited control reuse and under overlap weighting, the REM minus NREM heart-rate difference was 1.17 bpm (0.67 to 1.66) and 1.16 bpm (0.71 to 1.61), and REM sleep heart rate rose by 5.71 and 5.99 bpm (Supplementary Figure S4a). Redrawing the match in 500 bootstrap replicates, all of which produced a match, gave intervals of 4.35 to 7.71 bpm for REM sleep heart rate and 0.63 to 1.59 bpm for the REM minus NREM difference (Supplementary Figure S4b). The rematched difference averaged 1.13 bpm, close to the alternative designs, so the primary estimate of 1.42 bpm lies toward the upper end of the plausible range. Among starters of any GLP-1 receptor agonist, the difference was 1.03 bpm (0.68 to 1.38) under overlap weighting; among starters of liraglutide, dulaglutide or tirzepatide it was not distinguishable from zero (+0.57 bpm, −0.30 to 1.48, in 30 pairs), in a match that failed the balance check (Supplementary Figure S5).

Because heart rate during REM sleep starts higher than during NREM sleep, a single proportional rise would by itself produce a larger absolute increase during REM sleep. On the relative scale, however, the proportional heart-rate rise during REM sleep still exceeded that of NREM sleep (ratio 1.020, 95% CI 1.012 to 1.027; Figure 3b). The DiDs did not depend on the initiators’ baseline heart rate once regression to the mean was removed (Oldham slopes compatible with zero, p=0.25 to 0.90; Figure 3c). The REM minus NREM difference did not grow with each pair’s heart-rate change during nocturnal wake (slope −0.039, p=0.14) and was 1.58 bpm (1.06 to 2.09) when that change was zero (Figure 3d), so the extra REM increase adds to the general rise rather than scaling with it. Individual pairs varied widely (from −4.83 to +6.68 bpm, with 24 of 90 below zero; Figure 3a), so the larger REM increase describes the cohort average. REM sleep heart rate rose in every subgroup of sex, age, BMI and antihypertensive therapy, and the REM minus NREM difference was positive in each, with no subgroup difference distinguishable from zero (p=0.19 to 0.50; Supplementary Table S3). These analyses were chosen after the primary result, and their p values do not account for that choice.

**Figure 3.**
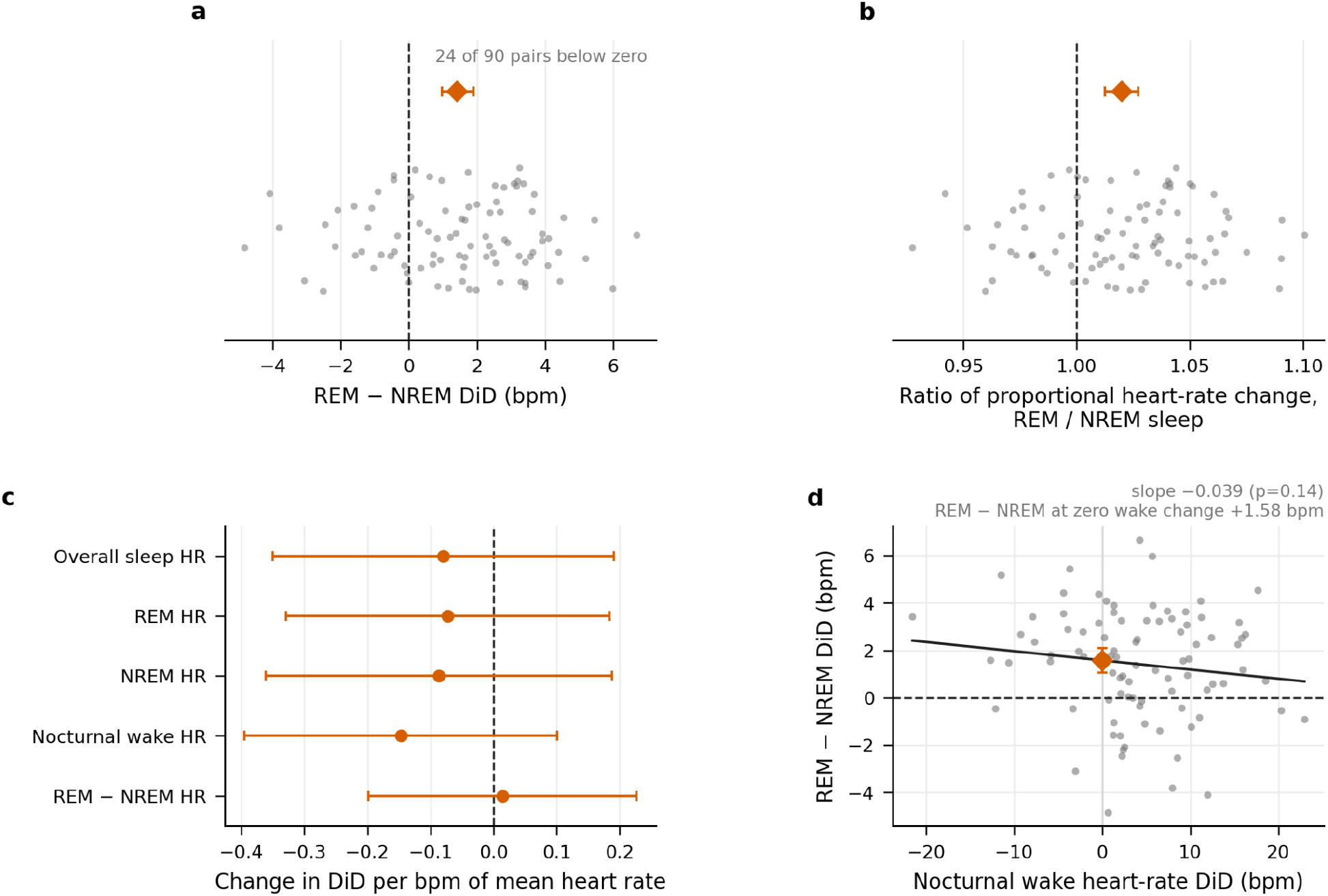
The larger REM increase is not explained by proportional scaling, baseline heart rate or the heart-rate change during nocturnal wake. (a) REM minus NREM heart-rate DiD for each matched pair (gray, 90 pairs) and the cohort estimate with its 95% CI (vermilion). (b) The same pairs as the ratio of the proportional heart-rate change in REM sleep to that in NREM sleep; 1.0 (dashed) is a single proportional rise; vermilion, geometric mean. (c) Oldham slope of each DiD on the initiator’s mean heart rate across the two visits (vermilion), with 95% CIs. (d) Each pair’s REM minus NREM heart-rate difference against its nocturnal wake DiD, with the fitted line (black) and the estimate at zero nocturnal wake change (vermilion). All four analyses are post hoc. HR, heart rate. *Alt text: Four panels in two rows. Panel a: a jittered scatter of 90 matched-pair REM minus NREM heart-rate differences, from minus 4.83 to plus 6.68 bpm, 24 of them left of a dashed zero line, with a vermilion diamond above the cloud at plus 1.42. Panel b: the same 90 pairs as ratios of proportional heart-rate change in REM to NREM sleep, from about 0.93 to 1.10, with a dashed line at 1.0 and a vermilion diamond at 1.020, 1.012 to 1.027. Panel c: for sleep overall, REM sleep, NREM sleep, nocturnal wake and the REM minus NREM contrast, a vermilion slope on mean heart rate between about minus 0.15 and plus 0.01, every interval spanning a dashed zero line. Panel d: a scatter of per-pair REM minus NREM difference against the nocturnal wake difference, from about minus 22 to plus 23 bpm, with a nearly flat fitted line and a vermilion diamond at zero nocturnal wake change showing 1.58 bpm, 1.06 to 2.09*.

### The larger REM increase is not an artifact of sleep-stage estimation

Because the device estimates sleep stage partly from pulse signals, a drug that raises heart rate could in principle change which periods are classified as REM sleep. Three observations argue against this. First, sleep-stage proportions across 16,417 HPP visits with more than three hours of sleep matched polysomnographic norms: REM sleep occupied 23.7% of sleep time (interquartile range 18.8 to 28.9), deep sleep 17.2% (13.7 to 20.9) and light sleep 58.3% (51.4 to 65.7). Second, in the matched pairs, semaglutide did not change the proportion of sleep time estimated as REM (DiD +0.85 percentage points, −0.98 to 2.69), and the REM minus NREM heart-rate difference did not track each pair’s change in that proportion. Third, the per-second recordings, which separate light from deep sleep, showed heart rate rising in every sleep stage, a larger rise during REM sleep than during light sleep (+0.81 bpm, 0.23 to 1.38) and no difference between deep and light sleep (+0.14 bpm, −0.59 to 0.90) (Supplementary Figure S6). This per-second match did not pass the balance check (maximum SMD 0.260), so it is descriptive, but it shows that a change in the mix of light and deep sleep cannot account for the larger REM increase.

### Weight loss and heart-rate variability do not explain the increase

Two further questions bear on how the heart-rate increase should be interpreted: whether it follows from weight loss, and whether the accompanying HRV decreases indicate a separate autonomic effect. Adjusting for each pair’s weight change left the REM sleep heart-rate increase unchanged (+6.25 bpm, 4.37 to 8.12, in 90 pairs), and neither REM sleep heart rate nor the REM minus NREM difference changed with time on drug (slopes −0.15 and −0.06 bpm per month; p=0.18 and 0.23). Weight loss without the drug went the other way. Among 5,053 participants who never took a GLP-1 drug, REM sleep heart rate fell by 0.14 bpm per kg of weight lost (95% CI 0.11 to 0.18), whereas in initiators it did not depend on weight change (−0.03 bpm per kg lost, −0.21 to +0.15; Figure 4a). Matched to participants who never took a GLP-1 drug and lost a similar amount of weight (−4.1 versus −4.4 kg; 90 pairs), initiators’ REM sleep heart rate rose by 5.88 bpm (4.67 to 7.11) and that of their matches by 0.40 bpm (−0.63 to 1.50), a difference of 5.48 bpm (3.80 to 7.20); matched on DXA fat loss (78 pairs), the changes were 5.32 and 0.05 bpm, a difference of 5.27 bpm (3.54 to 7.08) (Figure 4b). These comparisons were added after the primary result, and participants who lose weight on their own may differ in diet, activity and health. Four HRV outcomes decreased (pNN20 and SDNN during REM and NREM sleep; Figure 2a), but time-domain HRV depends arithmetically on heart rate, so a decrease can restate a heart-rate increase rather than add to it [24]. After adjustment for the simultaneous heart-rate change in the same sleep stage, every HRV estimate was compatible with zero (Supplementary Figure S7). The data therefore support a heart-rate effect, but not an independent reduction in vagal activity.

**Figure 4.**
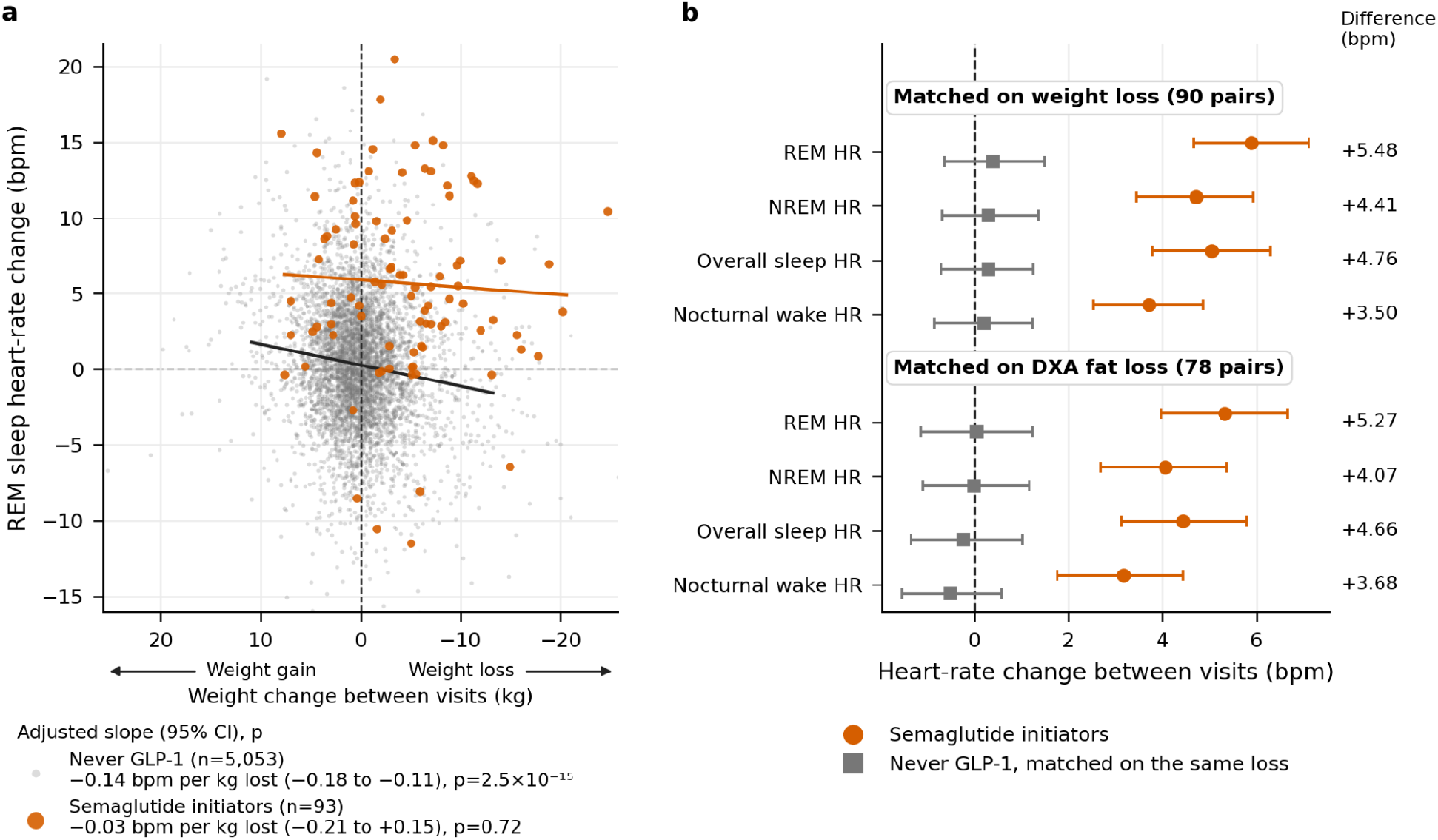
The heart-rate increase is not explained by weight loss. (a) Change in REM sleep heart rate against weight change between visits in participants who never took a GLP-1 drug (gray) and semaglutide initiators (vermilion), with least-squares lines; weight loss (negative change) is to the right of zero and weight gain to the left; slopes and their p values in the legend are adjusted for age, sex, baseline BMI and the interval between visits. (b) Heart-rate change between visits in initiators (vermilion circles) and in participants who never took a GLP-1 drug matched to them on the size of their weight loss (90 pairs) or DXA fat loss (78 pairs) (gray squares), with 95% CIs; numbers at right are the differences. Both panels are post hoc. HR, heart rate. *Alt text: Two panels. Panel a: a scatter of REM sleep heart-rate change against weight change, centered on zero with arrows marking weight gain to the left and weight loss to the right, with about 5,000 gray points for participants who never took a GLP-1 drug, whose fitted line falls slightly from left to right so that weight loss, on the right, goes with a small fall in heart rate, and 93 vermilion points for initiators, whose fitted line is nearly flat at about plus 5 bpm. Panel b: for REM sleep, NREM sleep, sleep overall and nocturnal wake, in two blocks matched on weight loss and on DXA fat loss, a vermilion circle for the initiators’ heart-rate change, between about plus 3 and plus 6 bpm, beside a gray square for their matches, near zero with intervals spanning it; the differences at right lie between plus 3.5 and plus 5.5 bpm*.

## Discussion

In this longitudinal cohort, semaglutide initiation was associated with lower blood pressure and body fat and with higher nocturnal heart rate. The principal extension of existing evidence is localization within sleep: heart rate rose during nocturnal wake, NREM sleep and REM sleep, and rose most during REM sleep, on a common matched sample. The larger heart-rate increase during REM sleep persisted under alternative matching designs, when the match was redrawn in a bootstrap and on a relative scale.

The result should be interpreted as a refinement of the known heart-rate effect of GLP-1 receptor agonists, not its discovery. Randomized and wearable studies already show that these drugs increase mean, resting, daytime and nighttime heart rate [4,9,10]. Our data add sleep-stage resolution and formal within-pair contrasts. The device-reported nightly summaries combine light and deep sleep into NREM sleep; the device’s per-second recordings, available for a subset, suggest no difference in the heart-rate increase between them.

Two features of that prior work bear on how the present magnitudes should be read. First, the comparison depends on when heart rate is measured. Pooled randomized data on clinic resting heart rate give smaller increases than we observe overnight: +1.86 bpm across the GLP-1 class [4] and +3.35 bpm for subcutaneous semaglutide in overweight or obesity without diabetes [25]. The one comparable ambulatory study, using 24-hour Holter recording, reported +6.3 bpm at night [9]. Our estimates for sleep overall (+5.18 bpm in the primary match and +5.15 bpm under overlap weighting) lie between these, and our nocturnal wake estimates (+3.89 and +4.03 bpm) are close to the clinic figure for semaglutide. Second, that ambulatory study found the increase larger at night than during the day (+6.3 versus +5.7 bpm). The present data show the same ordering, with the increase during sleep overall exceeding the nocturnal wake increase under both designs, consistent in direction with the only prior recording that separated day from night, although with a different agent, population and instrument.

Whether an increase of this size carries prognostic weight is not answerable here, but three lines of evidence explain why its timing matters. First, heart rate during sleep carries prognostic information of its own: resting heart rate is associated with cardiovascular and all-cause mortality in a graded fashion [6,7], and in ambulatory recordings a higher heart rate at night and a blunted nocturnal fall in heart rate are associated with cardiovascular events and mortality [26,27]. Because the overnight increases here exceed the clinic estimates, clinic measurements may understate the part of the class heart-rate effect that falls in this period. Second, REM sleep is the sleep stage in which surges in sympathetic activity are linked to nocturnal ischemia and arrhythmia in people with heart disease [28], and in trials of liraglutide in heart failure with reduced ejection fraction the drug raised heart rate without improving clinical stability or left ventricular function, with numerically more cardiac adverse events [29,30]. A heart-rate increase concentrated in REM sleep would therefore matter most for patients with coronary disease, arrhythmia or limited cardiac reserve, who were rare in this cohort. Third, an increase of 5 to 6 bpm is large enough to appear in the sleeping heart rate that consumer wearables report [10], where it is better read as an expected drug effect than as a sign of illness or poor recovery.

These associations are observational, none was established for heart rate within sleep stages, and the outcome trials that measured this class’s heart-rate effect still showed net cardiovascular benefit [2].

Several mechanisms could produce a larger heart-rate increase during REM sleep. A direct sinus-node effect, altered sympathovagal balance, changes in baroreflex regulation or central modulation that differs by sleep stage are all plausible; GLP-1 receptors are expressed at the sinoatrial node and at central autonomic sites, and both routes have been proposed for the class heart-rate effect [5,8]. REM sleep is characterized by sympathetic activity exceeding quiet wakefulness and by episodic surges in heart rate [11,12], so a sleep stage that is itself sympathetically driven could amplify a steady heart-rate effect. Our data locate the effect without identifying its route, and the additive behavior of the extra REM increase (Figure 3d) fits a stage-specific component at least as well as amplification. Measured weight change did not explain the REM sleep heart-rate increase, and weight loss without the drug went with a small fall in heart rate during sleep rather than a rise, so the increase is unlikely to follow from weight loss itself; because people who lose weight on their own are not a randomized comparison, this supports rather than proves that conclusion. Likewise, the disappearance of the HRV decreases after adjustment for heart rate argues against a separate vagal mechanism; it does not exclude subtle autonomic effects that would require beat-level data and dedicated physiological experiments.

The clinical findings support internal validity. The blood-pressure and body-composition changes agree in direction with randomized trials [1,31]; lean mass also fell (1.33 kg alongside 2.35 kg of fat mass), as reported in randomized body-composition studies [31]. Weight loss could be estimated only under overlap weighting, because the 1:1 anthropometric match failed balance, and its smaller magnitude than in high-dose obesity trials is compatible with heterogeneous real-world dose, duration, indication and adherence; trial weight loss is dose-dependent and substantially reversed on withdrawal [32]. The sitting-pulse contrast is generated mainly by a decline in the control group that the untreated cohort shared, so it is not an independent replication of the trial-observed rise in resting heart rate. The respiratory improvements (AHI −3.93 events/h) agree with randomized incretin trials in which liraglutide and tirzepatide reduced the AHI [33,34]. The diet estimates depended on overlap weighting after the 1:1 match failed balance, and gut taxonomy and microbiome function passed their matching criteria but produced no discovery; the diet changes are hypothesis-generating and are not developed here.

The study has several strengths. It uses within-person change over time, excludes every participant with a GLP-1 record from the controls, uses no control more than once in the primary design and makes retention and balance prerequisites for interpretation. Several estimators target related but distinct populations, and the central result survives explicit uncertainty in the matching step. Home sleep monitoring with sleep-stage estimation enables a comparison that is not normally available in trials or health records. More broadly, the study shows how a deeply phenotyped longitudinal cohort can emulate a drug trial for outcomes that trials have not measured, with the effects already known from randomized trials as a check on the design.

The limitations are equally important. Medication exposure is self-reported; dispensing, dose, adherence, indication and titration are unavailable, and unrecorded non-adherence or discontinuation would bias the estimates toward zero. The study is observational, and residual confounding remains possible despite close balance; we report no formal bound on how strong an unmeasured confounder would have to be to explain the contrast [35], and such an analysis would strengthen a replication. The DiD also assumes that, without the drug, initiators’ heart rate would have changed like their controls’; with one visit before the start this cannot be tested directly, although the matched controls changed like the whole untreated cohort (Supplementary Figure S2b). The sleep sample is modest, the non-semaglutide agents were too few and too imbalanced for a class-wide conclusion, and no external sleep cohort is included. Only 96 of the 494 initiators had paired sleep studies; they resembled the others in age, BMI and diabetes, but were less often female (49% vs 60%) and had started about a year earlier, as expected because recent starters had not yet returned for a follow-up visit (Supplementary Table S4). Participants were generally healthy adults aged 40 to 70 years in Israel who volunteered for a deep-phenotyping study, and few had diabetes (5% of the matched sleep cohort), so the estimates may not transfer directly to older patients, to people with type 2 diabetes or established cardiovascular disease, or to other populations. Pre-treatment heart rate was not matched on, by design, but differed between groups by less than 1 bpm, and the DiDs did not depend on baseline heart rate. REM sleep heart rate and the REM minus NREM contrast were designated after an exploratory analysis, so their inferential values should be regarded as internally validated rather than prospectively confirmed.

The WatchPAT does not record electroencephalography. It estimates sleep stage from PAT, pulse rate and actigraphy and has been validated against polysomnography [16,17], but agreement for individual periods is imperfect, and no participant here underwent simultaneous polysomnography. Sleep-stage proportions matched polysomnographic norms and the proportion of REM sleep did not change with treatment, which excludes a gross shift in stage estimation but not misclassification of individual periods, which could itself differ by sleep stage. A subset with concurrent polysomnography would settle this and is the most valuable addition a replication could make. Finally, the device-reported nightly summaries provide heart rate during REM sleep, NREM sleep and nocturnal wake; light and deep sleep are separated only in the per-second recordings, available at both visits for 60 matched initiators, and the HRV analyses use device-derived summaries rather than a raw electrocardiogram.

In conclusion, semaglutide initiation was associated with a heart-rate increase in every sleep stage, with a larger increase during REM sleep than during NREM sleep or nocturnal wake. Prospective replication with dispensed exposure, dose, polysomnographic sleep staging and an independent cohort is needed to determine the clinical and mechanistic significance of the larger heart-rate increase during REM sleep.

## Acknowledgments

We thank the members of the Segal group for fruitful discussions.

Use of artificial intelligence: Claude agent (Anthropic) was used to assist in writing the analysis code and in editing the manuscript text. All AI-assisted code was reviewed, tested, and validated by the authors, and all AI-assisted text was reviewed and revised by the authors, who take full responsibility for the content of this article. ChatGPT (OpenAI) was used to draw the study-design schematic in Figure 1a, which contains no data; the authors checked it against the design described in the Methods. The AI tools do not qualify for and are not listed as authors.

## Author contributions

Conceptualization: D.K., E.S. Data curation: D.K., S.K., A.Z., Y.S., A.G., Y.T.-B. Formal analysis: D.K., S.K., A.Z., Y.S., A.G., Y.T.-B. Funding acquisition: E.S. Investigation: D.K. Methodology: D.K., E.S. Project administration: E.S. Resources: E.S. Software: D.K. Supervision: E.S. Visualization: D.K., S.K., A.Z., Y.S., A.G., Y.T.-B. Writing – original draft: D.K. Writing – review & editing: D.K., S.K., A.Z., Y.S., A.G., Y.T.-B., E.S.

## Data availability

In accordance with institutional review board and privacy regulations, all datasets within the Human Phenotype Project (HPP) can be requested by qualified academic researchers at https://humanphenotypeproject.org/data-access, following the data sharing protocols established in the baseline cohort report [13]. Interested investigators should direct requests to to complete a non-commercial data-use agreement, after which access is provided within a secure cloud analysis platform.

## Code availability

The analysis code is available at https://github.com/kronga/glp1-rem-sleep-hr.

## Supplementary information

### Supplementary methods

#### Exposure ascertainment

Medication records carry a free-text name and a start date. A record was classified as semaglutide if its name matched the generic name or a brand name (Ozempic, Wegovy or Rybelsus, in English or Hebrew), and as another GLP-1 receptor agonist if it matched liraglutide (Saxenda, Victoza), dulaglutide (Trulicity) or tirzepatide (Mounjaro). The screen for other agents used every medication row, including rows without a start date, so a participant with any record of another agent was excluded from the semaglutide cohort, and a participant with any GLP-1 receptor agonist record was excluded from the control pool. A conservative floor on the start date (2018 for semaglutide) removed plainly impossible dates. Participants who switched between agents were not pooled into the primary cohort.

#### Visit selection

Visits were ordered by their measurement dates rather than by visit label. For each initiator and modality, the baseline visit was the last visit with a usable measurement strictly before the recorded start date and the follow-up visit the first usable visit at least three months after it, so that every baseline visit precedes the start date and every follow-up visit follows it. Controls could contribute any baseline and follow-up visit pair that an initiator used, and each control was matched within the same pair of visits.

#### Covariates

Visceral adipose tissue area was imputed within groups defined by sex and visit when it was not measured, and a missing-measurement indicator entered the propensity model. Continuous covariates were standardized before logistic-regression propensity estimation. Age, sex, BMI and waist and hip circumference were required for matching.

#### Overlap weighting

Overlap weights were one minus the propensity score for initiators and the propensity score for controls [1]. They were estimated separately within each exact-matching stratum (baseline visit, follow-up visit, sex and lipid-lowering therapy), so that the weighted comparison respects the same strata as the 1:1 match, and they were recalibrated within each stratum to the participants with each outcome available. Repeated eligible visit windows of a control were collapsed to that participant, and standard errors were clustered by participant.

#### Per-second sleep recordings

For the light and deep sleep comparison and for the heart rate around REM onset, heart rate and sleep stage were read from the per-second device recordings. The per-second analysis kept the primary cohort’s baseline and follow-up visits and matched controls again, by the same procedure, among participants whose per-second recordings were usable at both visits. For the REM-onset curves, each REM episode lasting at least three minutes was aligned on its onset, heart rate was averaged across episodes and nights for each participant and visit, and pairs with a usable curve at both visits in both members were kept.

#### Measurement protocols

Weight, height, waist and hip circumferences, sitting blood pressure and pulse, and whole-body DXA were measured at study visits. DXA used a Lunar Prodigy Advance scanner with CoreScan software (GE Healthcare), which estimates visceral and subcutaneous adipose tissue within the android region [2]. Blood tests were taken from participants’ health maintenance organization records. Stool was self-collected at home with OMNIgene·GUT OMR-200 kits (DNA Genotek) and shotgun-sequenced; species abundances were estimated by mapping reads to the cohort’s reference of gut species genomes [3], and pathway abundances with HUMAnN3 [4]. Diet was self-logged in the study smartphone app for about two weeks around each visit [4]; each log was assigned to the nearest visit within 60 days, and a visit needed at least three logged days. Home sleep monitoring used the WatchPAT-300 on up to three nights within two weeks; respiratory events were detected and sleep stages estimated by the device algorithm, with hypopneas requiring at least 3% oxygen desaturation for the AHI and at least 4% for the 4%-desaturation AHI, and desaturations of at least 4% for the ODI [4]. Medications, diagnoses including diabetes, and smoking were self-reported in online questionnaires and the study app, and diagnoses were reviewed with research staff at the visit [2].

### Supplementary results

#### Cohort flow

Supplementary Figure S1 traces both arms of the sleep analysis. Of 603 participants with any semaglutide record, 75 had a record of another GLP-1 receptor agonist and 34 had no start date or one before 2018, leaving 494 initiators. Of these, 96 had a usable sleep study before the recorded start and another at least three months after it. The other 398 had no usable sleep study at any visit (131, including participants with no dated study visit), none before the start (73), none after it (174) or a follow-up study less than three months after the start (20). Ninety-five had complete matching covariates and at least one sleep outcome, 94 were matched, and 90 had REM and NREM heart rate at both visits: one participant was lost to incomplete data, one to the propensity caliper and four to a missing REM or NREM heart-rate value. On the control side, 642 of the 13,714 HPP participants with a dated study visit had a GLP-1 receptor agonist record. Of the remaining 13,072, 5,503 had sleep studies at both visits of an initiator’s visit pair and complete matching covariates, and 94 of them were matched. Initiators with and without paired sleep studies are compared in Supplementary Table S4.

**Supplementary Figure S1.**
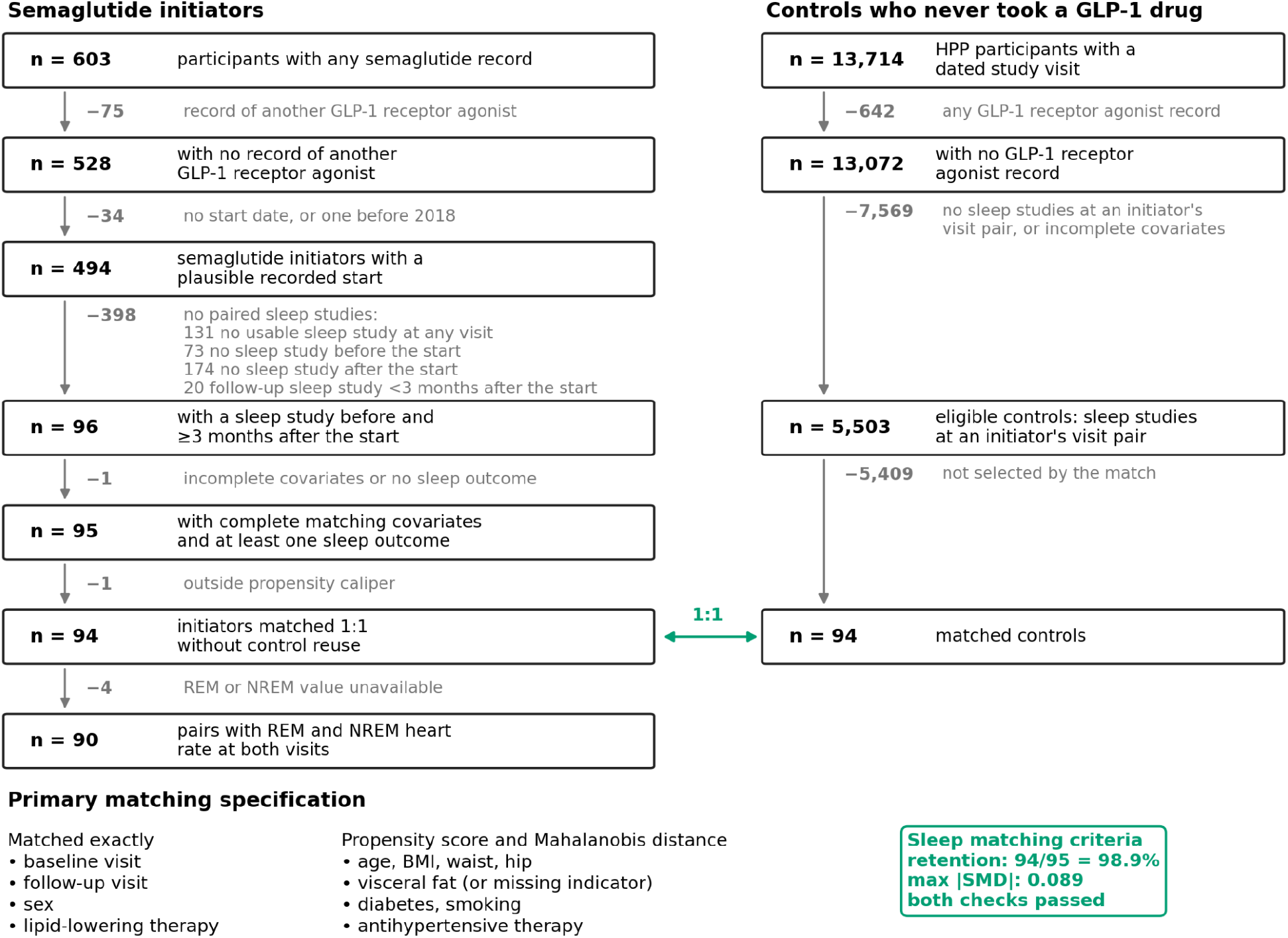
Cohort flow and matching specification for the sleep analysis. Left, semaglutide initiators; right, controls who never took a GLP-1 drug. Boxes give the participants remaining at each step and arrows the number removed and the reason; the green arrow marks the 1:1 match. Bottom, the exact-matching variables and propensity covariates; the green box reports the sleep matching criteria. *Alt text: A two-column flow chart. Left column, initiators, with a downward arrow between boxes carrying the number removed and the reason: 603 participants with any semaglutide record; minus 75 with a record of another GLP-1 receptor agonist, leaving 528; minus 34 with no start date or one before 2018, leaving 494; minus 398 without paired sleep studies (131 with no usable sleep study at any visit, 73 with none before the start, 174 with none after it, 20 with a follow-up study less than three months after the start), leaving 96; minus 1 with incomplete data, leaving 95; minus 1 outside the propensity caliper, leaving 94 matched; minus 4 without a REM or NREM value, leaving 90. Right column, controls: 13,714 HPP participants with a dated study visit; minus 642 with any GLP-1 receptor agonist record, leaving 13,072; minus 7,569 without sleep studies at an initiator’s visit pair or with incomplete covariates, leaving 5,503 eligible controls; minus 5,409 not selected, leaving 94 matched controls, joined to the 94 matched initiators by a green two-headed arrow labeled 1:1. Below, lists of the exact-matching variables and the propensity covariates beside a green box reporting 98.9% retention and a maximum absolute SMD of 0.089*.

#### All outcomes in the modalities that passed their matching criteria

Supplementary Table S1. lists every outcome in the sleep, blood-pressure, DXA and gut-taxonomy modalities, with its DiD, within-modality q and discovery status. The 272 microbial pathways of the microbiome-function modality, none of which was a discovery, are not listed.

**Supplementary Table S1.**
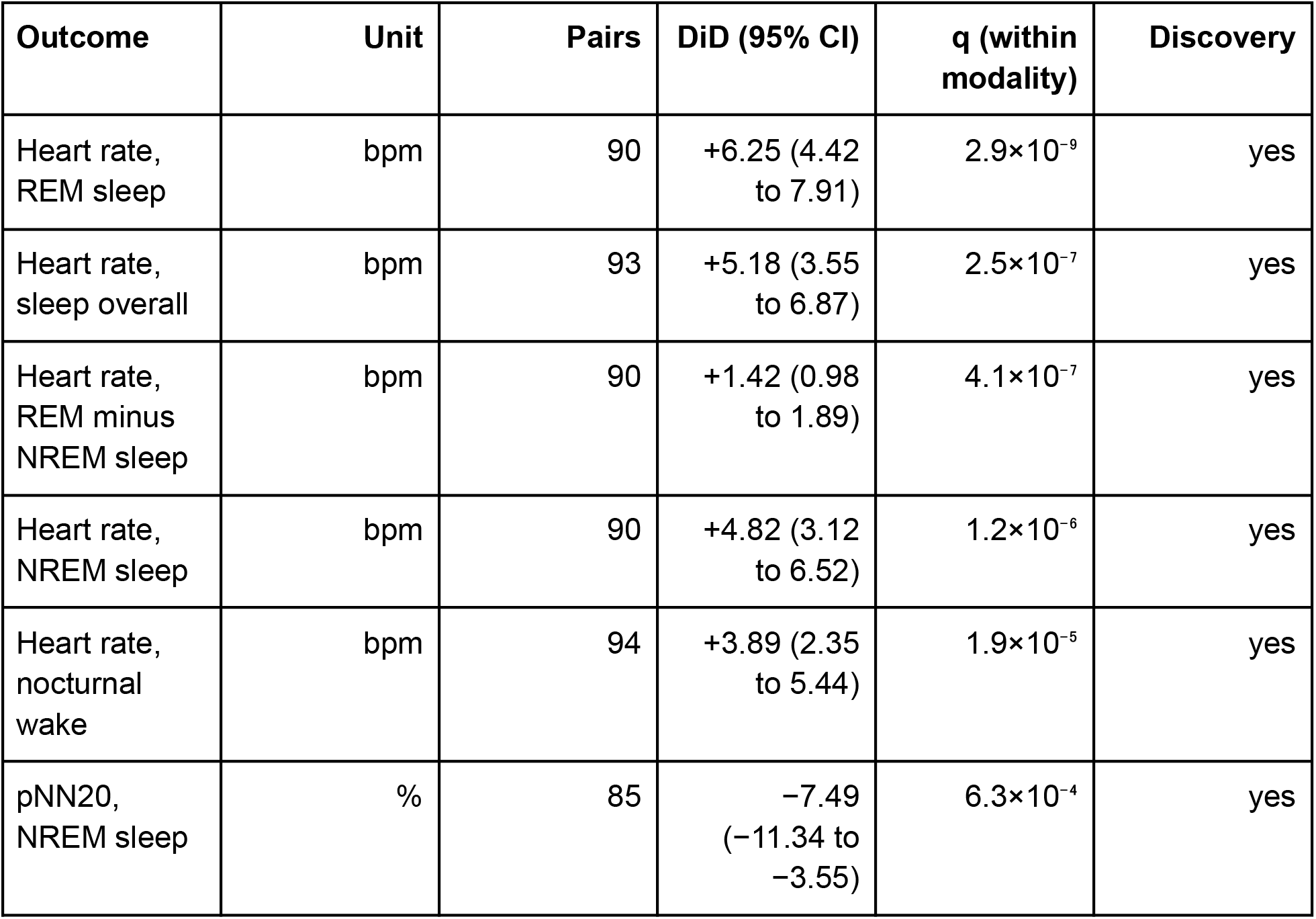
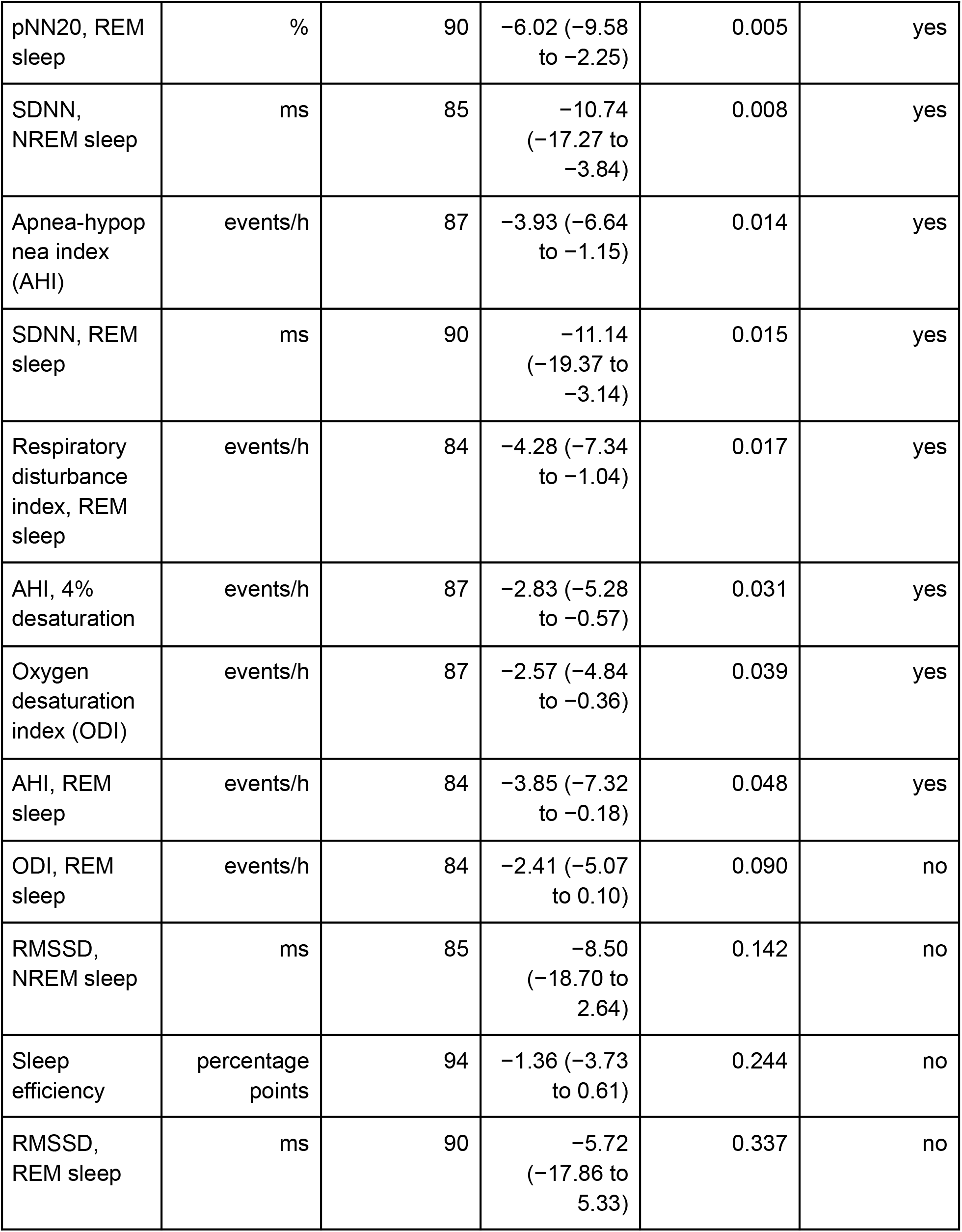
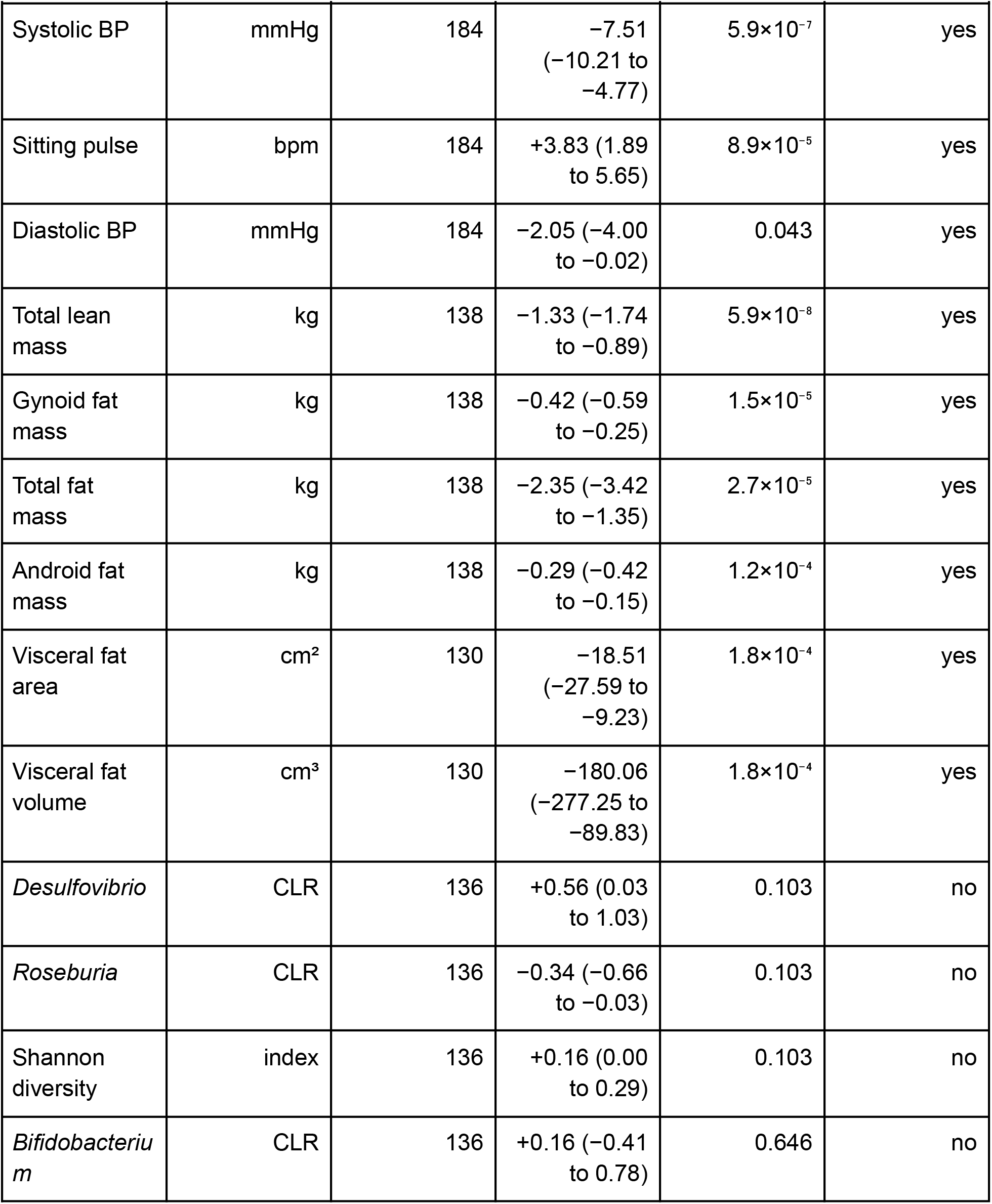
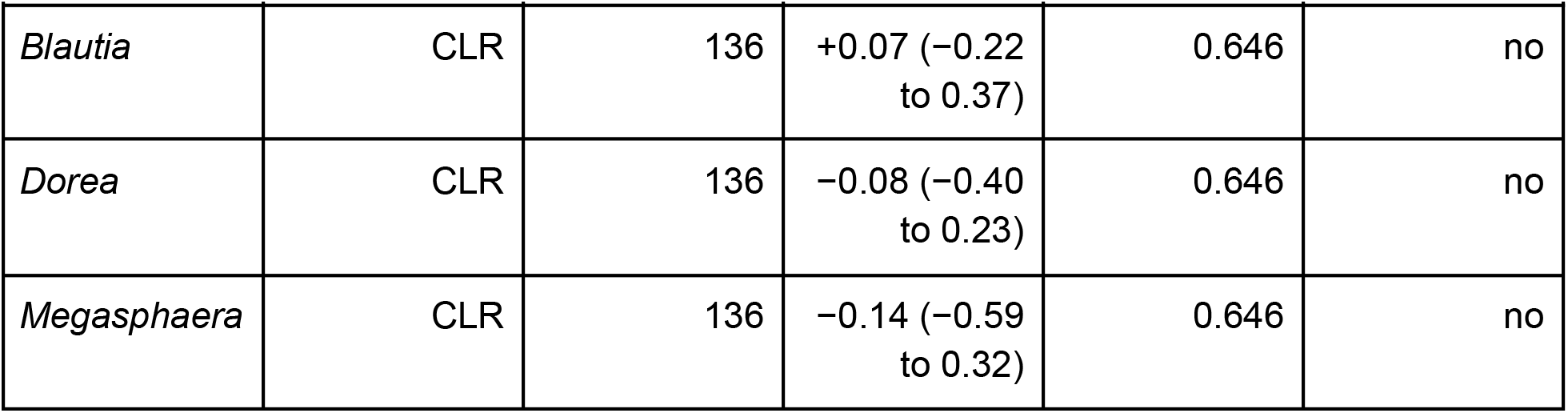
Outcomes in the sleep, blood-pressure, DXA and gut-taxonomy modalities. DiD with 95% bootstrap CIs. q is the Benjamini-Hochberg value within the modality; a discovery also required its modality to pass the hierarchical modality test. Heart-rate variability is RMSSD, SDNN and pNN20; respiratory indices are events per hour; DXA masses are in kilograms; gut taxa are centered log-ratio (CLR) abundances. The gut-taxonomy modality passed its matching criteria but was not selected by the modality-level test, so none of its outcomes is a discovery.

| Outcome | Unit | Pairs | DiD (95% CI) | q (within modality) | Discovery |
| --- | --- | --- | --- | --- | --- |
| Heart rate, REM sleep | bpm | 90 | +6.25 (4.42 to 7.91) | $2.9 \times 10^{-9}$ | yes |
| Heart rate, sleep overall | bpm | 93 | +5.18 (3.55 to 6.87) | $2.5 \times 10^{-7}$ | yes |
| Heart rate, REM minus NREM sleep | bpm | 90 | +1.42 (0.98 to 1.89) | $4.1 \times 10^{-7}$ | yes |
| Heart rate, NREM sleep | bpm | 90 | +4.82 (3.12 to 6.52) | $1.2 \times 10^{-6}$ | yes |
| Heart rate, nocturnal wake | bpm | 94 | +3.89 (2.35 to 5.44) | $1.9 \times 10^{-5}$ | yes |
| pNN20, NREM sleep | % | 85 | -7.49 (-11.34 to -3.55) | $6.3 \times 10^{-4}$ | yes |

| **Outcome** | **Unit** | **Pairs** | **DiD (95% CI)** | **q (within modality)** | **Discovery** |
| --- | --- | --- | --- | --- | --- |
| pNN20, REM sleep | % | 90 | -6.02 (-9.58 to -2.25) | 0.005 | yes |
| SDNN, NREM sleep | ms | 85 | -10.74 (-17.27 to -3.84) | 0.008 | yes |
| Apnea-hypopnea index (AHI) | events/h | 87 | -3.93 (-6.64 to -1.15) | 0.014 | yes |
| SDNN, REM sleep | ms | 90 | -11.14 (-19.37 to -3.14) | 0.015 | yes |
| Respiratory disturbance index, REM sleep | events/h | 84 | -4.28 (-7.34 to -1.04) | 0.017 | yes |
| AHI, 4% desaturation | events/h | 87 | -2.83 (-5.28 to -0.57) | 0.031 | yes |
| Oxygen desaturation index (ODI) | events/h | 87 | -2.57 (-4.84 to -0.36) | 0.039 | yes |
| AHI, REM sleep | events/h | 84 | -3.85 (-7.32 to -0.18) | 0.048 | yes |
| ODI, REM sleep | events/h | 84 | -2.41 (-5.07 to 0.10) | 0.090 | no |
| RMSSD, NREM sleep | ms | 85 | -8.50 (-18.70 to 2.64) | 0.142 | no |
| Sleep efficiency | percentage points | 94 | -1.36 (-3.73 to 0.61) | 0.244 | no |
| RMSSD, REM sleep | ms | 90 | -5.72 (-17.86 to 5.33) | 0.337 | no |
| Systolic BP | mmHg | 184 | -7.51<br>(-10.21 to -4.77) | $5.9 \times 10^{-7}$ | yes |
| Sitting pulse | bpm | 184 | +3.83 (1.89 to 5.65) | $8.9 \times 10^{-5}$ | yes |
| Diastolic BP | mmHg | 184 | -2.05 (-4.00 to -0.02) | 0.043 | yes |
| Total lean mass | kg | 138 | -1.33 (-1.74 to -0.89) | $5.9 \times 10^{-8}$ | yes |
| Gynoid fat mass | kg | 138 | -0.42 (-0.59 to -0.25) | $1.5 \times 10^{-5}$ | yes |
| Total fat mass | kg | 138 | -2.35 (-3.42 to -1.35) | $2.7 \times 10^{-5}$ | yes |
| Android fat mass | kg | 138 | -0.29 (-0.42 to -0.15) | $1.2 \times 10^{-4}$ | yes |
| Visceral fat area | cm<sup>2</sup> | 130 | -18.51<br>(-27.59 to -9.23) | $1.8 \times 10^{-4}$ | yes |
| Visceral fat volume | cm<sup>3</sup> | 130 | -180.06<br>(-277.25 to -89.83) | $1.8 \times 10^{-4}$ | yes |
| *Desulfovibrio* | CLR | 136 | +0.56 (0.03 to 1.03) | 0.103 | no |
| *Roseburia* | CLR | 136 | -0.34 (-0.66 to -0.03) | 0.103 | no |
| Shannon diversity | index | 136 | +0.16 (0.00 to 0.29) | 0.103 | no |
| *Bifidobacterium* | CLR | 136 | +0.16 (-0.41 to 0.78) | 0.646 | no |
| *Blautia* | CLR | 136 | +0.07 (−0.22 to 0.37) | 0.646 | no |
| *Dorea* | CLR | 136 | −0.08 (−0.40 to 0.23) | 0.646 | no |
| *Megasphaera* | CLR | 136 | −0.14 (−0.59 to 0.32) | 0.646 | no |

#### Change in each group and in untreated participants

Each DiD combines an initiator change and a control change (Supplementary Figure S2a). The nocturnal heart-rate result comes from the initiators: REM sleep heart rate rose by 5.52 bpm in initiators and fell by 0.72 bpm in controls. The sitting-pulse contrast comes mostly from the controls: initiators rose by 1.58 bpm while controls fell by 2.24 bpm. Controls also fell by 3.51 mmHg in diastolic pressure.

A control group can change for reasons specific to its members rather than because untreated people change over the same interval. Supplementary Figure S2b therefore compares the matched controls with every eligible participant who never took a GLP-1 drug, 5,401 to 7,782 people depending on the outcome, reweighted to the matched pairs’ visit intervals. The untreated-cohort change lies inside the matched controls’ interval for nine of the ten outcomes. For lean mass the matched controls lost 0.33 kg (0.04 to 0.62) against 0.03 kg in the untreated cohort, which if anything makes the lean-mass DiD conservative. For diastolic pressure the untreated cohort itself fell by 4.33 mmHg, and for sitting pulse by 1.80 bpm; both are read from the same seated clinic measurement, which fits a repeat-visit effect better than aging. The DiD removes this change either way, and it does not bear on the nocturnal heart-rate result.

**Supplementary Figure S2.**
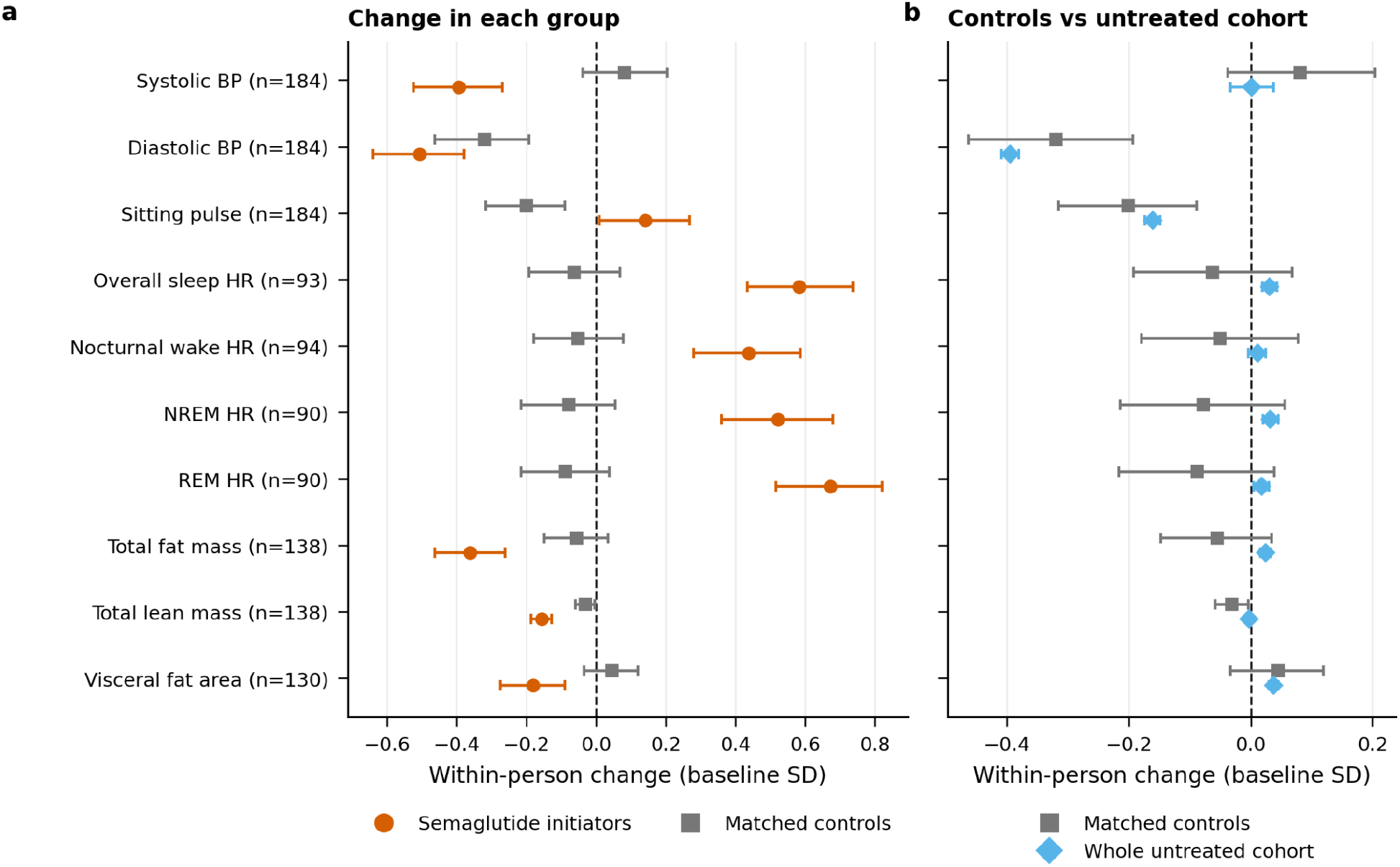
Change in each group, and matched controls against the untreated cohort. (a) Within-person change in initiators (vermilion circles) and matched controls (gray squares), in baseline standard deviations with 95% bootstrap CIs; the DiD is the distance between them. (b) Matched-control change (gray squares) against the change in all eligible participants who never took a GLP-1 drug (light blue diamonds) over the same visit intervals. HR, heart rate; BP, blood pressure. *Alt text: Two panels sharing ten rows: systolic, diastolic and sitting pulse; heart rate during sleep overall, nocturnal wake, NREM and REM sleep; and DXA total fat mass, lean mass and visceral fat area. Panel a shows a vermilion circle for initiators and a gray square for controls in each row; initiators’ blood pressure and body fat fall and their sleep heart rate rises by about 0.45 to 0.7 SD while controls stay near zero; for sitting pulse initiators rise slightly and controls fall. Panel b shows the gray control squares beside light blue diamonds for the untreated cohort, whose intervals are very narrow; every diamond except that for lean mass lies inside the corresponding control interval*.

#### Matching criteria by modality

Every modality was matched and checked before any outcome was examined (Supplementary Figure S3; Supplementary Table S2). Sleep, blood pressure, DXA body composition, gut taxonomy and microbiome function passed both the retention and the balance checks, and anthropometrics, blood tests and diet failed the balance check. Passing the checks permits inference but does not imply a discovery: gut taxonomy and microbiome function passed and produced none.

**Supplementary Figure S3.**
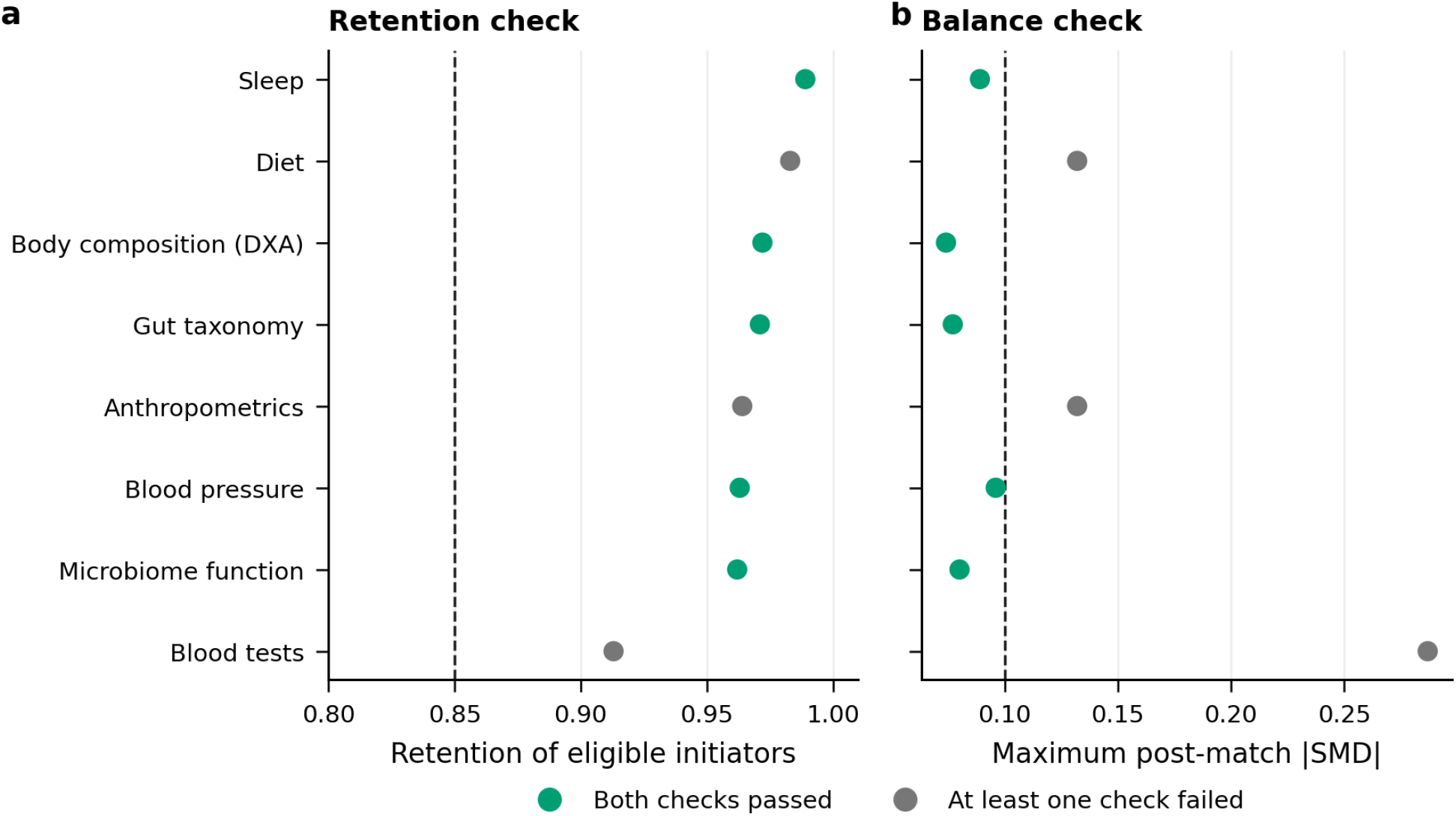
Retention and balance checks for every modality. (a) Fraction of eligible initiators retained by 1:1 matching; dashed line, 85% threshold. (b) Maximum post-match absolute SMD; dashed line, 0.10. Green, passed both checks; gray, failed. *Alt text: Two dot plots over eight modalities. Panel a shows retention against a dashed line at 0.85; every modality exceeds it. Panel b shows maximum post-match absolute SMD against a dashed line at 0.10. Green dots, which passed both checks, are sleep, blood pressure, body composition, gut taxonomy and microbiome function; gray dots are anthropometrics and diet just above 0.10 and blood tests near 0.3*.

**Supplementary Table S2.**
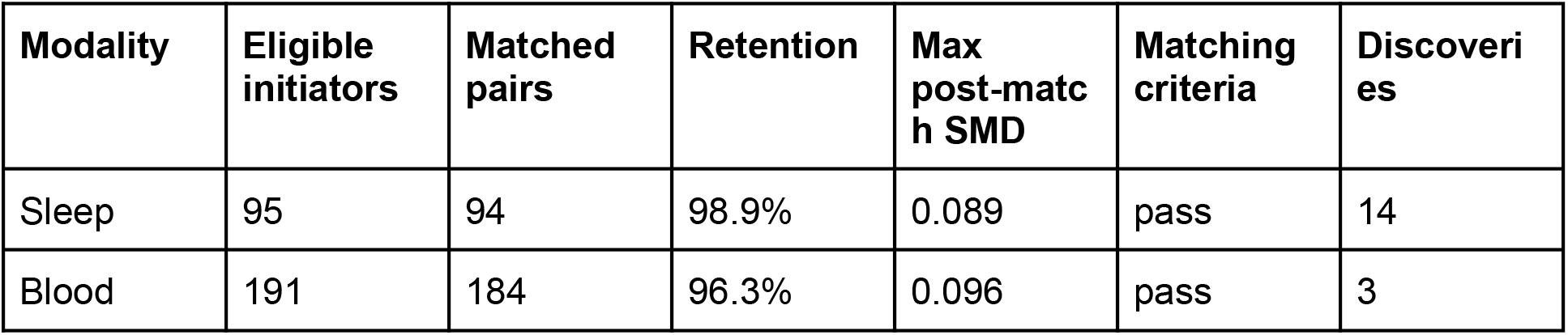
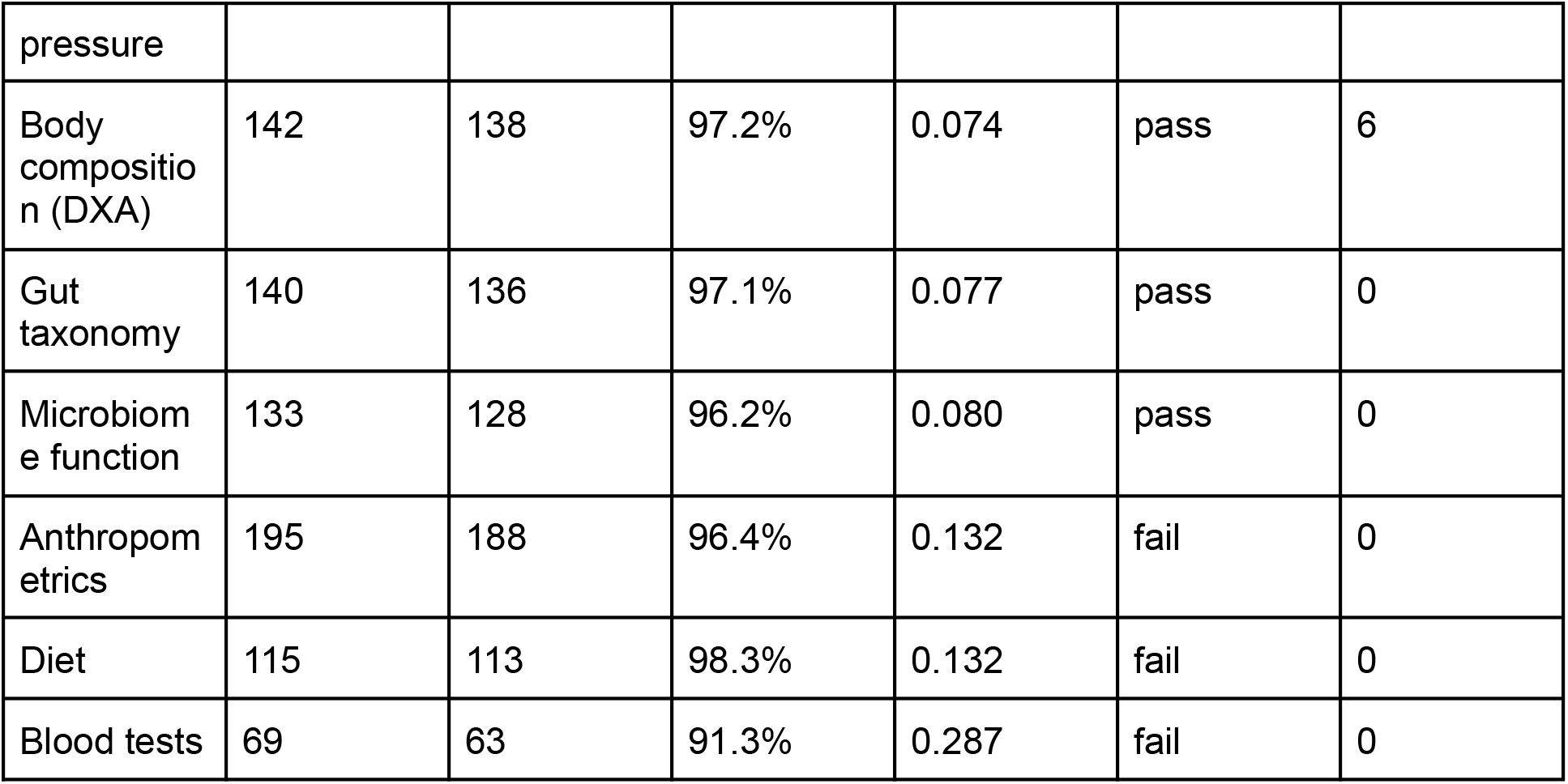
Matched design by modality. A modality was interpreted if matching retained at least 85% of eligible initiators and the maximum post-match absolute SMD was below 0.10. Discoveries are outcomes that passed the hierarchical false-discovery procedure.

#### Alternative designs and rematching

The main estimates condition on a single 1:1 match without control reuse. Supplementary Figure S4a repeats five of them under 1:1 to 1:3 matching with limited control reuse and under overlap weighting. REM sleep heart rate rose by 6.25, 5.71 and 5.99 bpm, and the REM minus NREM heart-rate difference was 1.42, 1.17 and 1.16 bpm, under the primary, limited-reuse and overlap designs, all of which met the sleep matching criteria with every interval above zero. Weight (−3.53, −2.92 and −3.11 kg), systolic pressure (−7.51, −6.77 and −6.14 mmHg) and sitting pulse (+3.83, +3.33 and +3.79 bpm) behaved the same way.

Bootstrapping the matched pairs does not include the uncertainty of the matching step itself. Supplementary Figure S4b therefore refits the propensity model and redraws the match in each of 500 bootstrap replicates, all of which produced a match. The rematching intervals are about as wide as the matched-pair ones, and every one excludes zero: 4.35 to 7.71 bpm for REM sleep heart rate against 4.42 to 7.91, and 0.63 to 1.59 bpm for the REM minus NREM difference against 0.98 to 1.89. The rematched difference averaged 1.13 bpm, below the observed 1.42 and close to the alternative designs, so 1.42 bpm is better read as the upper part of a 1.1 to 1.4 bpm range than as a precise value.

**Supplementary Figure S4.**
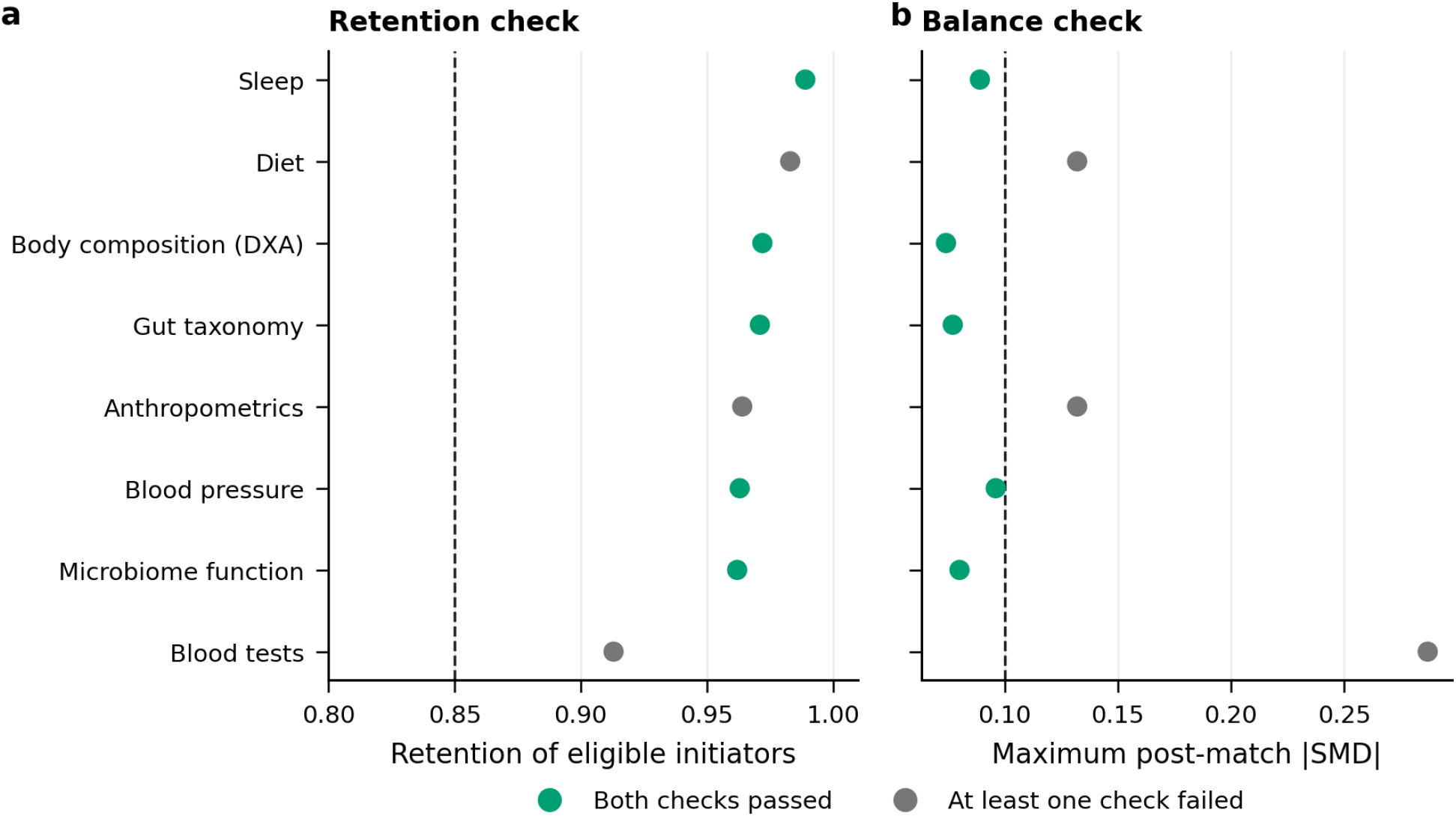
Alternative matching designs and rematching. (a) Five outcomes in baseline standard deviations under the primary 1:1 match (vermilion circles), 1:1 to 1:3 matching with each control used at most twice (green squares) and overlap weighting (pink diamonds); open markers failed the matching criteria. (b) Five sleep heart-rate outcomes in bpm with the matched-pair bootstrap interval (vermilion) and the interval when the match is redrawn in 500 replicates (green), both at the observed estimate. HR, heart rate; BP, blood pressure. *Alt text: Two panels. Panel a: five outcomes, each shown three times in baseline standard deviations, as a vermilion circle for the primary match, a green square for limited reuse and a pink diamond for overlap weighting, with open markers for failed matching criteria; within every outcome the three estimates sit close together on the same side of zero. Panel b: five sleep heart-rate outcomes, sleep overall, nocturnal wake, NREM sleep, REM sleep and the REM minus NREM difference, each with a vermilion matched-pair interval and a green rematching interval; all exclude zero, and for the REM minus NREM difference the rematching interval, 0.63 to 1.59 bpm, sits lower than the matched-pair interval, 0.98 to 1.89*.

#### Exposure-definition sensitivity

The same procedure was repeated for two alternative exposure definitions (Supplementary Figure S5). Among the 739 starters of any GLP-1 receptor agonist, a superset of the primary cohort, weight and systolic blood pressure fell with passing 1:1 matches (−2.80 kg and −6.16 mmHg). The sleep match in this group failed the balance check (maximum SMD 0.130); under overlap weighting, which passed, REM sleep heart rate rose by 4.62 bpm (3.57 to 5.66) and the REM minus NREM heart-rate difference was 1.03 bpm (0.68 to 1.38) in 152 initiators. Both are smaller than in the semaglutide cohort, as expected if the added agents contribute less. Among the 170 starters of liraglutide, dulaglutide or tirzepatide, every design failed the balance check (maximum SMD 0.20 to 0.35) and the sleep analysis rested on 30 to 31 initiators. Weight fell in this group as well, by 3.82 kg (1.41 to 6.46), but neither the REM sleep heart-rate increase (+1.86 bpm, −0.82 to 4.60) nor the REM minus NREM difference (+0.57 bpm, −0.30 to 1.48) was distinguishable from zero. This group cannot separate a smaller agent-specific effect from imprecision and residual imbalance.

**Supplementary Figure S5.**
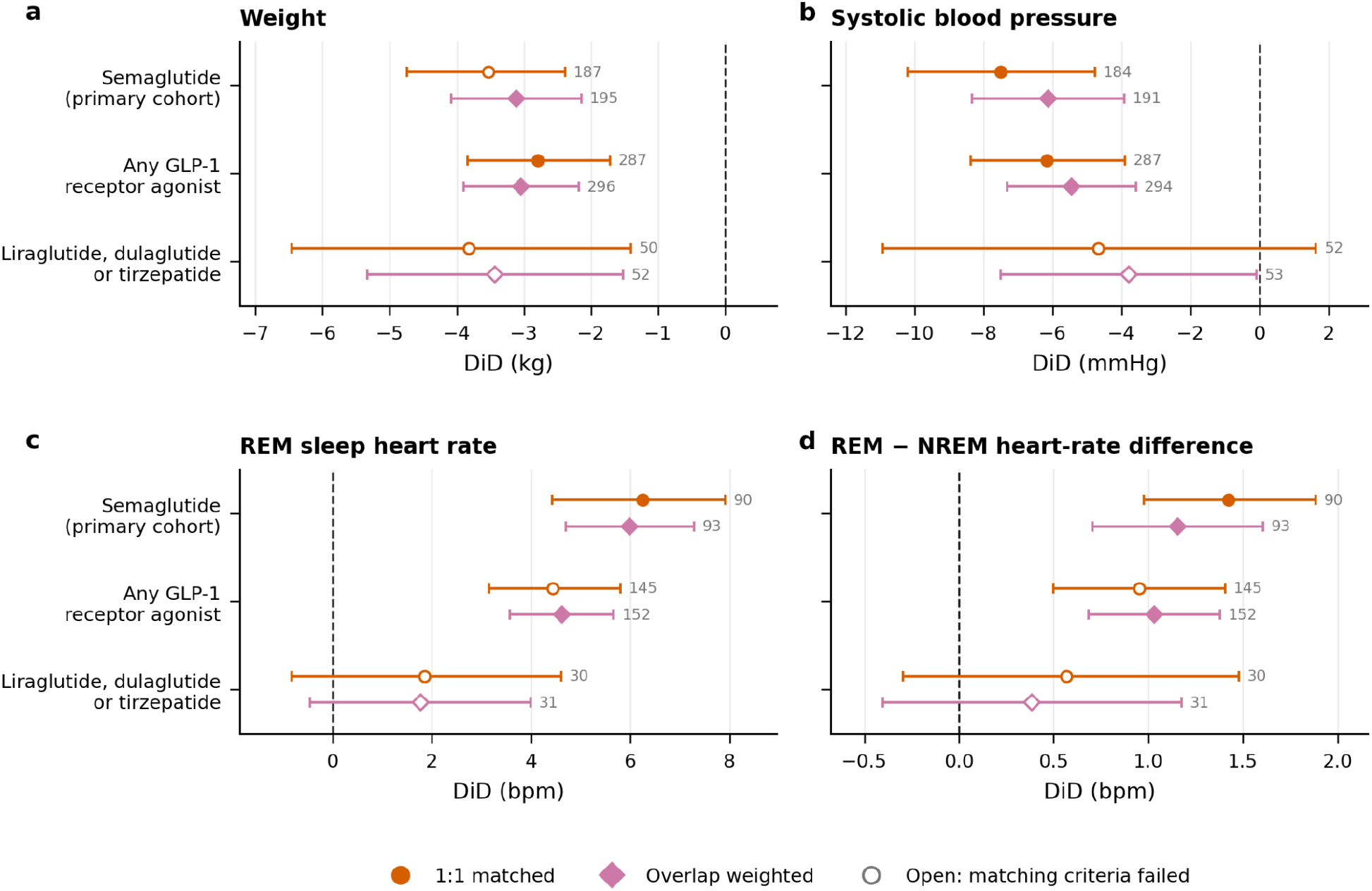
Exposure-definition sensitivity. DiD for (a) weight, (b) systolic blood pressure, (c) REM sleep heart rate and (d) the REM minus NREM heart-rate difference in the primary semaglutide cohort (494 initiators), starters of any GLP-1 receptor agonist (739) and starters of liraglutide, dulaglutide or tirzepatide (170). Vermilion circles, 1:1 matched; pink diamonds, overlap weighted; open markers failed the matching criteria. Numbers give matched pairs or contributing initiators. *Alt text: Four forest plots, for weight, systolic blood pressure, REM sleep heart rate and the REM minus NREM heart-rate difference, sharing three rows: semaglutide, any GLP-1 receptor agonist, and liraglutide, dulaglutide or tirzepatide. Each row has a vermilion circle for the 1:1 match and a pink diamond for overlap weighting, filled when the matching criteria were met. Semaglutide and any-GLP-1 estimates agree in direction and exclude zero in every panel, with smaller any-GLP-1 sleep estimates. The liraglutide, dulaglutide or tirzepatide row is open throughout; its weight estimates lie below zero, while its REM heart-rate and contrast intervals, on about 30 initiators, span zero*.

#### Light and deep sleep from the per-second recordings

The device-reported nightly summaries combine light and deep sleep into NREM sleep. The device’s per-second recordings separate them, so the primary cohort’s visits were matched again among participants with usable per-second recordings at both visits (Supplementary Figure S6). Sixty of 64 eligible initiators were matched, but the match failed the balance check (maximum SMD 0.260), so it is descriptive. Heart rate rose in every sleep stage: by 6.01 bpm during REM sleep (4.14 to 7.98), 5.20 bpm during light sleep (3.36 to 7.03), 5.34 bpm during deep sleep (3.29 to 7.39) and 4.40 bpm during nocturnal wake (2.38 to 6.42). The heart-rate increase during REM sleep exceeded that of NREM sleep by 0.80 bpm (0.19 to 1.39) and that of light sleep by 0.81 bpm (0.23 to 1.38); the REM minus deep difference was similar in size but imprecise (+0.67 bpm, −0.34 to 1.60), and deep and light sleep did not differ (+0.14 bpm, −0.59 to 0.90).

**Supplementary Figure S6.**
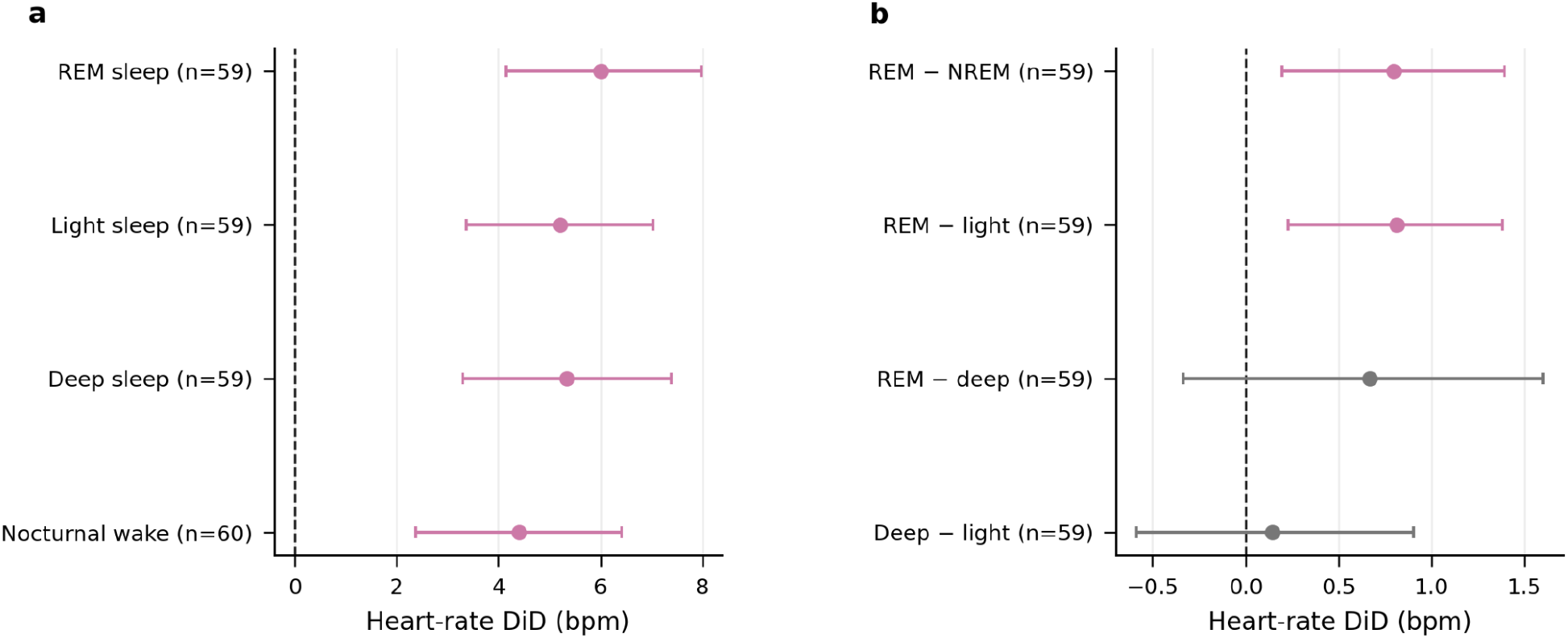
Heart rate by sleep stage from the per-second recordings. (a) Heart-rate DiD by sleep stage (59 pairs; nocturnal wake, 60). (b) Within-night heart-rate differences between sleep stages (59 pairs). Pink, 95% bootstrap interval excludes zero; gray, includes zero. Descriptive: this match failed the balance check. *Alt text: Two forest plots. Panel a: heart-rate DiDs by sleep stage, all positive with pink intervals excluding zero: REM sleep plus 6.01 bpm, light sleep plus 5.20 and deep sleep plus 5.34 on 59 pairs each, and nocturnal wake plus 4.40 on 60 pairs. Panel b: within-night heart-rate differences on 59 pairs; REM minus NREM, plus 0.80, and REM minus light, plus 0.81, are pink with intervals excluding zero, while REM minus deep, plus 0.67, and deep minus light, plus 0.14, are gray with intervals spanning zero*.

#### Heart-rate variability adjusted for heart rate

Four HRV outcomes were discoveries in the primary analysis: pNN20 and SDNN during REM and NREM sleep. Time-domain HRV depends arithmetically on heart rate, so each HRV DiD was also estimated with adjustment for the simultaneous heart-rate DiD in the same sleep stage (Supplementary Figure S7). Every adjusted estimate moved toward zero and every adjusted interval included zero, so the HRV decreases restate the heart-rate increase rather than add an independent reduction in vagal activity.

**Supplementary Figure S7.**
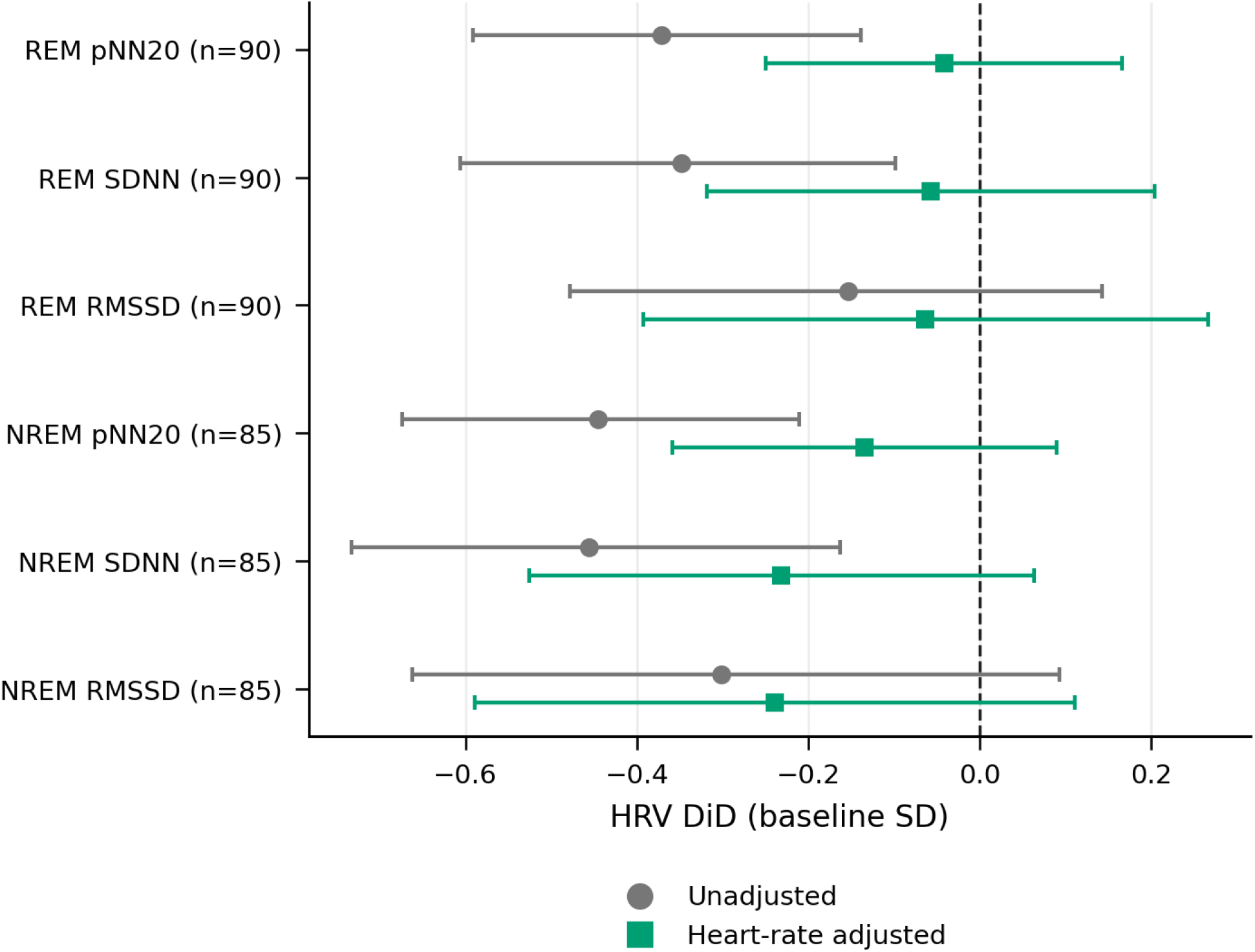
Heart-rate variability before and after adjustment for heart rate. HRV DiD in baseline standard deviations, unadjusted (gray circles) and adjusted for the simultaneous heart-rate change in the same sleep stage (green squares), with 95% CIs; 90 (REM sleep) and 85 (NREM sleep) pairs. *Alt text: A forest plot of six HRV outcomes, pNN20, SDNN and RMSSD during REM sleep (90 pairs) and NREM sleep (85 pairs), in baseline standard deviations. Each outcome appears twice: unadjusted as a gray circle and adjusted for the simultaneous heart-rate change as a green square. All unadjusted estimates are negative and four exclude zero: pNN20 and SDNN during REM and NREM sleep. Every adjusted estimate moves toward zero and every adjusted interval includes zero*.

#### Subgroups

The REM sleep heart-rate increase and the REM minus NREM difference were estimated within subgroups of the 90 primary pairs, defined by the initiator’s sex, age (below or at least the median of 51.2 years), BMI (below or at least 30 kg/m²) and antihypertensive therapy at the baseline visit (Supplementary Table S3). The subgroups were chosen after the primary analysis; diabetes was too rare (5 matched initiators) to examine. REM sleep heart rate rose in every subgroup, by 4.80 to 7.82 bpm, and the REM minus NREM difference was positive in every subgroup, from 0.93 to 1.65 bpm. No difference between subgroups was distinguishable from zero (p=0.19 to 0.50), although with 15 to 62 pairs per subgroup only large differences could have been detected.

**Supplementary Table S3.** REM sleep heart rate and the REM minus NREM difference by subgroup. DiD with 95% pair-bootstrap CIs in the 90 primary pairs, split by the initiator’s value at the baseline visit; sex was matched exactly. Difference is the second subgroup minus the first, with its 95% CI and p from a regression of the pair DiD on the subgroup indicator with HC3 standard errors. Post hoc and descriptive.

| Subgroup | Pairs | REM sleep heart rate, bpm | REM minus NREM heart rate, bpm |
| --- | --- | --- | --- |
| All pairs | 90 | +6.25 (4.42 to 7.91) | +1.42 (0.98 to 1.89) |
| Male | 43 | +6.99 (4.78 to 9.29) | +1.64 (0.88 to 2.41) |
| Female | 47 | +5.56 (3.21 to 7.92) | +1.23 (0.66 to 1.78) |
| Difference |  | -1.43 (-4.85 to 1.98);<br>p=0.41 | -0.41 (-1.38 to 0.56);<br>p=0.41 |
| Age < 51.2 years | 45 | +6.92 (4.25 to 9.52) | +1.61 (1.04 to 2.16) |
| Age ≥ 51.2 years | 45 | +5.58 (3.57 to 7.57) | +1.24 (0.50 to 1.97) |
| Difference |  | -1.34 (-4.77 to 2.09);<br>p=0.45 | -0.37 (-1.33 to 0.59);<br>p=0.45 |
| BMI < 30 kg/m<sup>2</sup> | 28 | +7.82 (5.18 to 10.62) | +0.93 (0.04 to 1.86) |
| BMI ≥ 30 kg/m<sup>2</sup> | 62 | +5.53 (3.36 to 7.59) | +1.65 (1.07 to 2.17) |
| Difference |  | -2.29 (-5.90 to 1.32);<br>p=0.22 | +0.72 (-0.35 to 1.79);<br>p=0.19 |
| No antihypertensive therapy | 75 | +6.54 (4.59 to 8.50) | +1.51 (1.02 to 1.97) |
| Antihypertensive therapy | 15 | +4.80 (2.35 to 7.33) | +0.98 (-0.41 to 2.36) |
| Difference |  | -1.74 (-5.08 to 1.60);<br>p=0.31 | -0.53 (-2.07 to 1.00);<br>p=0.50 |

#### Initiators with and without paired sleep studies

Of the 494 initiators, 96 had a sleep study before the recorded start and another at least three months after it. Baseline characteristics at the last visit before the start were available for all 96 and for 244 of the other 398 (Supplementary Table S4). The two groups were similar in age, BMI, diabetes and lipid-lowering therapy. Initiators with paired sleep studies were less often female (49.0% vs 60.2%), slightly more often on antihypertensive therapy (18.8% vs 11.9%) and had started earlier (mean 2023.3 vs 2024.3), as expected because recent starters had not yet returned for a follow-up visit.

**Supplementary Table S4.** Initiators with and without paired sleep studies. Values at the last visit before the recorded start with age, sex and BMI measured. Continuous variables are mean ± SD and binary variables n (%). SMD, standardized mean difference.

| Characteristic | With paired sleep studies (n=96) | Without paired sleep studies (n=244 of 398) | SMD |
| --- | --- | --- | --- |
| Age, years | 53.0 ± 7.8 | 52.0 ± 7.3 | 0.129 |
| Body-mass index, kg/m<sup>2</sup> | 31.6 ± 4.1 | 31.3 ± 3.9 | 0.094 |
| Waist circumference, cm | 102.4 ± 12.3 | 99.4 ± 13.6 | 0.234 |
| Year of recorded start | 2023.3 ± 1.2 | 2024.3 ± 1.4 | -0.747 |
| Female sex | 47 (49.0%) | 147 (60.2%) | -0.228 |
| Diabetes | 5 (5.2%) | 10 (4.1%) | 0.053 |
| Current smoking | 4 (4.2%) | 18 (7.4%) | -0.138 |
| Antihypertensive therapy | 18 (18.8%) | 29 (11.9%) | 0.191 |
| Lipid-lowering therapy | 10 (10.4%) | 33 (13.5%) | -0.096 |

